# The National COVID-19 Epi Model (NCEM): Estimating cases, admissions and deaths in South Africa

**DOI:** 10.1101/2022.09.05.22279174

**Authors:** Sheetal Prakash Silal, Juliet R.C. Pulliam, Gesine Meyer-Rath, Lise Jamieson, Brooke E Nichols, Jared Norman, Rachel Hounsell, Saadiyah Mayet, Frank Kagoro, Harry Moultrie

## Abstract

**Background:** In March 2020 the South African COVID-19 Modelling Consortium was formed to support government planning for COVID-19 cases and related healthcare. Models were developed jointly by local disease modelling groups to estimate cases, resource needs and deaths due to COVID-19.

**Methods:** The National COVID-19 Epi Model (NCEM) while initially developed as a deterministic compartmental model of SARS-Cov-2 transmission in the nine provinces of South Africa, was adapted several times over the course of the first wave of infection in response to emerging local data and changing needs of government. By the end of the first wave, the NCEM had developed into a stochastic, spatially-explicit compartmental transmission model to estimate the total and reported incidence of COVID-19 across the 52 districts of South Africa. The model adopted a generalised Susceptible-Exposed-Infectious-Removed structure that accounted for the clinical profile of SARS-COV-2 (asymptomatic, mild, severe and critical cases) and avenues of treatment access (outpatient, and hospitalisation in non-ICU and ICU wards).

**Results:** Between end-March and early September 2020, the model was updated several times to generate new sets of projections and scenario analyses to be shared with planners in the national and provincial Departments of Health, the National Treasury and other partners in a variety of formats such as presentations, reports and dashboards. Updates to model structure included finer spatial granularity, limited access to treatment, and the inclusion of behavioural heterogeneity in relation to the adoption of Public Health and Social Measures. These updates were made in response to local data and knowledge and the changing needs of the planners.

**Conclusions:** The NCEM attempted to incorporate a high level of local data to contextualise the model appropriately to address South Africa’s population and health system characteristics. Origin and contextualisation of data and understanding of the population’s interaction with the health system played a vital role in producing and updating estimates of resource needs, demonstrating the importance of harnessing and developing local modelling capacity.

## Background

On 31 December 2019, the World Health Organization (WHO) reported a cluster of pneumonia cases in Wuhan City, China known as COVID-19, the infectious disease caused by ‘Severe Acute Respiratory Syndrome Coronavirus 2’ (SARS-CoV-2)^1^. By November 2020, 216 countries and territories had reported over 47,000,000 cases and 1,200,000 deaths^2^. At the time in South Africa, at the end of the first wave of infection, COVID-19 cases were detected in all nine provinces reaching over 725,000 reported cases in the public and private sectors combined^3^.

Aimed at understanding the spread of the disease and the impacts of different interventions, a number of mathematical models of the burden and cost of the COVID-19 epidemic have been developed over the two years across low- and middle-income countries (LMIC). Mathematical models provide a valuable framework for analysing the transmission and impact of infectious diseases. The application of disease transmission models and costing tools provide a platform to turn the surveillance data collected by national control programmes into strategic information to support policy makers in programme and funding decisions.

In South Africa, the South African COVID-19 Modelling Consortium (SACMC), convened by the National Institute for Communicable Diseases (NICD) on behalf of the National Department of Health (NDOH), coordinated mathematical modelling efforts. This group of researchers from academic, non-profit, and government institutions was formed to provide, assess and validate model projections to support planning by the South African government. Two models were developed: the National COVID-19 Epi Model (NCEM) developed jointly by disease modelling groups at the University of Cape Town, Stellenbosch University, and the University of the Witwatersrand, and the National COVID-19 Cost Model (NCCM) developed by the group at the University of the Witwatersrand. These models have been used since the end of March 2020 to project cases, resource needs and deaths and assess the extent to which these were impacted by emerging variants^4-13^.

From the start of the pandemic, uncertainty existed in almost all central aspects of SARS-CoV-2, including its prevalence, transmission, the proportion of infected people who remain asymptomatic, the role of seasonality, whether cross-immunity to the virus from other infections exists, and the extent to which immunity to the virus itself persists - as well as the impact of any of the implemented Public Health and Social Measures (PHSM) such as social distancing and mask wearing. Since the emergence of variants in December 2020 and introduction of vaccination, additional uncertainty arose with respect to variant characteristics and vaccine effectiveness. Despite this uncertainty, governments around the world have had to draft and update policy in response to this changing global threat.

South Africa is in the unusual position among LMIC in that it has access to existing public health infrastructure such as surveillance networks and data collection systems, as well as local capacity for infectious disease modelling and economic analysis. The COVID-19 pandemic has brought to light the similarities in and differences between country approaches to epidemic decision-making and response. LMIC in particular have had to balance higher baseline disease burden from other causes with the potential for economic contraction and existing demands on the healthcare system. From a modelling perspective, the availability of data, demand for projections and the use of scenarios in determining health policy may vary between LMIC and High Income Countries.

The purpose of this paper is to present the NCEM and its features used to estimate cases, deaths and hospitalisations over the course of the first wave of the epidemic in South Africa for planning purposes by the South African government with a particular focus on demonstrating its rapid evolution in response to local data and knowledge. The NCEM was updated several times over the course of the first wave in South Africa as local data became available and the need for projections at different stages of the treatment pathway and at different spatial scales changed. In this period from March 2020 to October 2020, the primary use of the NCEM was for projection of the timecourse of case numbers and hospitalisations, as well as cumulative indicators such as the number of expected COVID-19-associated deaths. The modelling process, model evolution and a reflection on modelling for this purpose and in this context that could highlight best practices and lessons learned to assist LMIC with modelling for pandemic preparedness is presented in Meyer-Rath, Hounsell (14). This paper presents the technical modelling framework used for projection, the consideration of behavioural factors in updating the framework and its application at a district level for the first wave of COVID-19 in South Africa.

### Methodology

The National COVID-19 Epi Model (NCEM) was initially developed as a deterministic compartmental model of SARS-Cov-2 transmission in the nine provinces of South Africa. After observing case and death trends and model performance for a few months at the start of the pandemic, and in line with the requests for decision-making support at a finer spatial granularity, the NCEM underlying structure, assumptions and geographic scope was updated to a stochastic, compartmental, spatially-explicit transmission model to estimate the total and reported incidence of COVID-19 across the 52 districts of South Africa (structural updated model as at 5 September 2020). The updated model followed a generalised Susceptible-Exposed-Infectious-Removed (SEIR) structure that incorporated disease severity (asymptomatic, mild, severe and critical cases) and access to treatment in and out of hospital (outpatients, Intensive Care Unit (ICU) and non-ICU care) (Figure 1).

**Figure 1.**
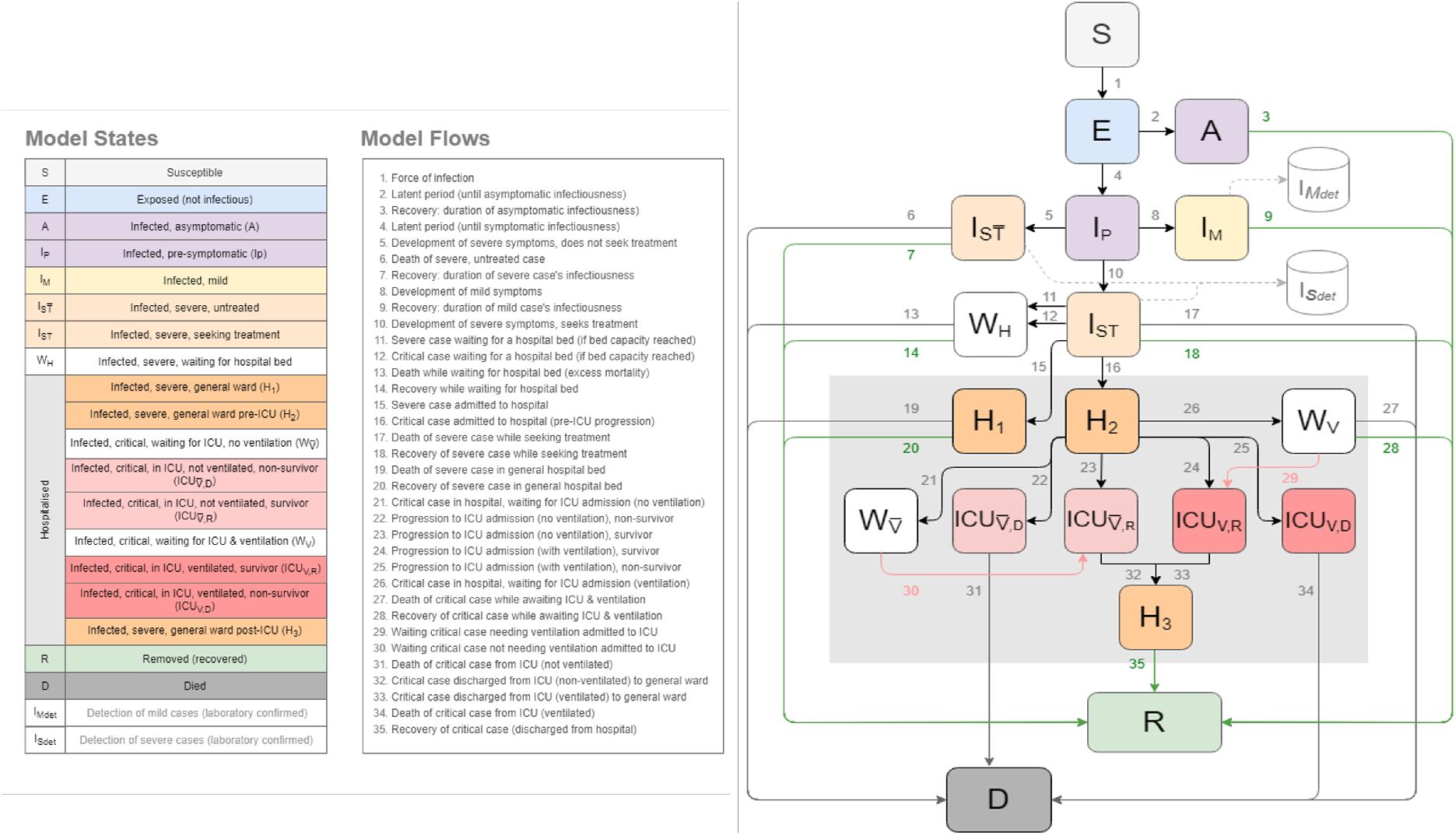
The National COVID-19 Epi Model structure (updated)^7^

Key features of the updated NCEM include:

### Hospital treatment pathway

The pathway in hospital was modelled such that patients may be treated in a non-ICU (general medical) or an ICU ward, with or without support by mechanical ventilation in ICU. Informed by admissions data, the pathway was divided into a series of compartments to allow for different times to recovery and death. To incorporate limited capacity, a series of ‘waiting’ compartments was modelled to simulate individuals who, due to capacity constraints, are in need of a hospital or ICU bed but unable to occupy one. As beds become available, patients in these waiting compartments may transition to general hospital/ ICU compartments.

### Treatment seeking

behaviour and health access was modelled such that not all severely and critically ill individuals are assumed to access hospital-level care. Individuals in these compartments experience higher mortality rates than those who receive the appropriate level of care (e.g., patients who require but do not receive mechanical ventilation are assumed to have 100% mortality).

### Detection

Given that many infected individuals are asymptomatic or mildly symptomatic and are not likely to seek out a diagnostic test, and additionally, owing to the severe laboratory capacity constraints in South Africa, not all infected individuals will be diagnosed. Changing priorities in testing of hospitalised patients and non-hospitalised mild cases over the course of the epidemic were captured in the model.

### Spatial scale

The scale of the model is at the district level (subnational administrative level two), reflecting the population size and connectivity of each of the 52 districts in South Africa. Baseline movement between districts is estimated based on aggregate cell phone mobility data from 1 March 2020. District-to-district connectivity matrices were constructed based on the proportion of mobile phone events that occur in each district outside the home district. The home district is defined as the place where a mobile device is located between 10 pm and 4 am. Separate matrices were constructed for each lockdown restriction to reflect the average levels of movement between districts for each period.

### Behavioural heterogeneity

was incorporated through an adjustment to the force of infection. This captures the phenomenon that differences exist in contact patterns among individuals where some individuals experience different risks and exhibit different behavioural patterns, with highly connected individuals becoming infected earlier in the epidemic and infecting more contacts. The force of infection (transmission function) was altered to decrease as immunity builds up in the most connected individuals early on in the epidemic^15^.

The model was developed in R and C++, with the deterministic differential equations solved using the deSolve package in R^16, 17^, and stochastic differential equations solved using the diffeqr package in Julia^18^. A parallel implementation is also available in the code.

The update to NCEM was informed by a scenario analysis that considered alternate behavioural and epidemiological assumptions and are described in detail in the Results section and Supplementary file.

### Data

Case and hospital admissions and deaths data were obtained from NICD’s Notifiable Medical Conditions database and Data for COVID (DATCOV), the sentinel hospital surveillance database respectively.

Contact rates for the population were based on analyses on the reproductive number over time^19, 20^ and Google mobility data Google COVID-19 Community Mobility Reports^21^ while district level population data for 2020 was obtained from Statistics South Africa^22^. Mobility between districts over time was estimated from mobile phone event data from Vodacom South Africa where daily event data (excluding static devices) with district source and destination information were normalised to establish the proportion of daily events per source in each destination district. This was used to generate average proportion of daily events matrices over the periods of restrictions. Total deaths due to COVID-19 were estimated from the South African Medical Research Council’s Weekly Excess Deaths reports^23, 24^.

Disaster-related restrictions in South Africa took the form of a series of alert levels that were gradually relaxed over the course of the epidemic^25^. These alert levels, generally implemented uniformly across all districts, enforced social distancing, restricted movement between districts, and reduced contacts among individuals, but also limited trade in different sectors over the course of the epidemic^26^. As the restrictions affected business sectors differentially, and representative local-level data on adherence to and impact of PHSM did not exist at the time, general measures were used to account for the impact of the alert levels on reducing average contacts in the population over time. Changes in average contacts were estimated from the effective reproductive number over time based on analyses by the NICD for early periods of restrictions (alert levels 5 and 4) when the impact of a build-up in population immunity was low^20^. For alert levels 3, 2 and 1 thereafter, changes in contact rates at the provincial level were updated based on reductions in time spent in places of residence as determined by Google Mobility Trend Data^21^.

## Results

### Original model performance

On 12 June 2020, we published a set of short-term projections to estimate cases and deaths for June/July^6^. We first estimated cumulative detected cases and cumulative deaths (right) from 21 March 2020 to 15 July 2020 (Figure 2). These projections show that the original NCEM model closely estimated the reported cumulative detected cases observed for the projection period. It was during this period that daily deaths and admissions began to flatten in the Western Cape province, for reasons that were not yet well understood at the time. These results were strongly caveated in the original report as not incorporating potentially important elements such as heterogeneity and behavioural response to the epidemic. Given that infectious disease models such as the NCEM are mechanistic models driven by the underlying biology of the virus, population-level behaviour characteristics, and the care pathways, without prior knowledge of how these factors would play out, it was not possible to quantitatively predict the deceleration in daily cases and deaths that would result.

**Figure 2.**
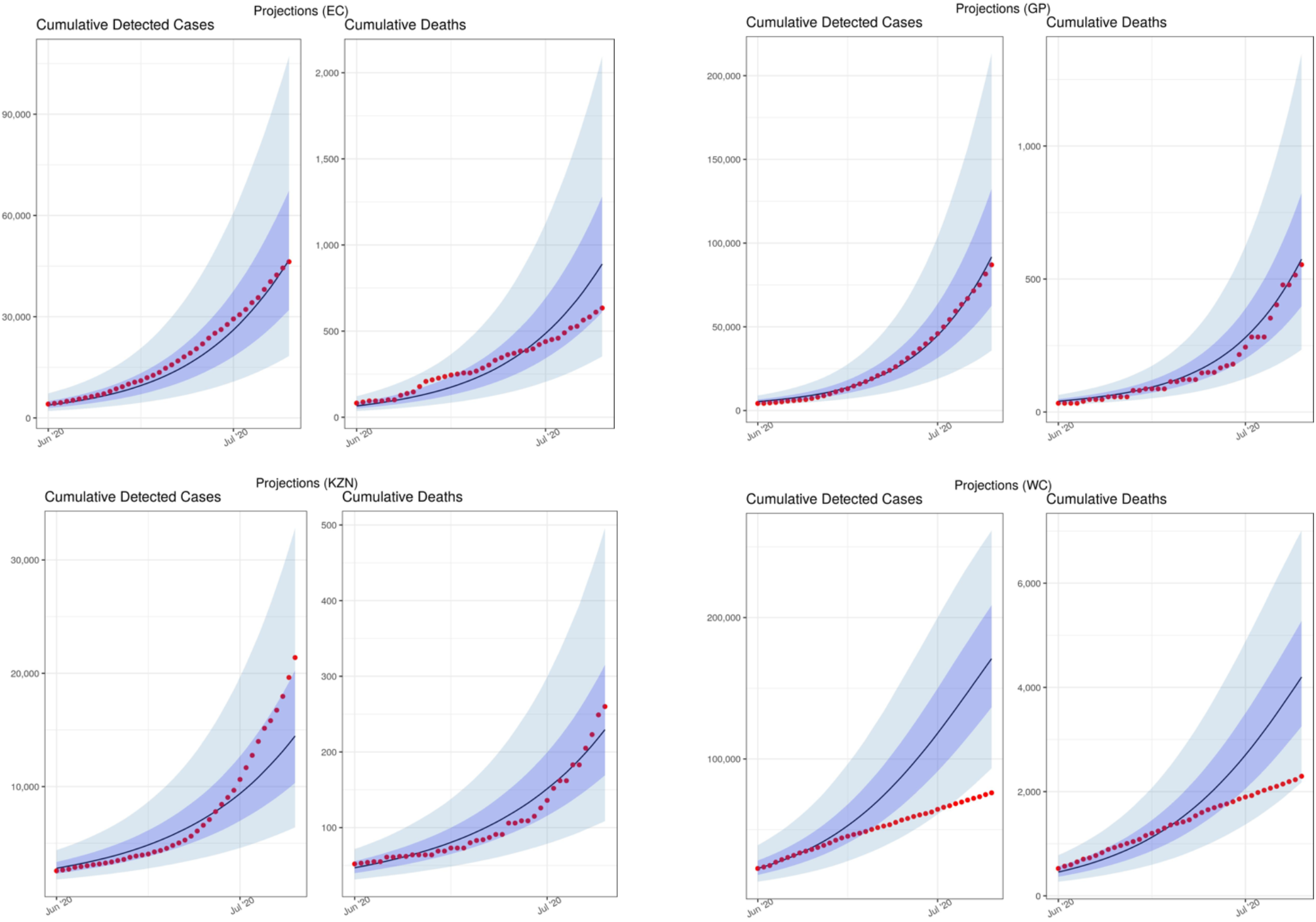
Model performance: Previously projected (12 June) vs observed cumulative detected cases and cumulative deaths (selected provinces: Eastern Cape (EC), Gauteng (GP), KwaZulu-Natal (KZN) and Western Cape (WC)) [median projected= black lines; observed data= red dots]

The deviation of the model from the observed trends in detected cases and reported deaths lead to additional uncertainty and confusion amongst policymakers and public as to the likely end of the wave and the implication for provinces where cases were still low. This also had a negative impact on the perceived credibility of the model where models tended to be viewed as forecasts, rather than tools generating projections under a set of assumptions. We responded to perform a number of scenario analyses to interrogate the impact that four distinct factors could have had in explaining the difference between the NCEM projections from June and reported case and death data in the Western Cape.

These were:

a. A **lower than assumed population attack rate**, possibly due to different levels of susceptibility in different population groups (including children) or the presence of existing T-cell derived-immunity after prior exposure to other coronaviruses. This is modelled by allowing a proportion of individuals to be immune throughout the course of the epidemic.
b. **Behaviour change in response to an increased local death rate**. This scenario takes into account a potential impact of public awareness of the increasing deaths and the looming threat of overwhelmed healthcare facilities in the Western Cape, which, combined with communication campaigns, may have resulted in better adherence to non-pharmaceutical interventions (NPIs) (e.g. masks, hand washing and physical distancing) and in those most at risk for severe COVID-19 disease taking additional precautions to isolate themselves. This is modelled by allowing the population in each district to reduce interactions when district death rates are high and increase interactions when death rates are low.
c. **Better adherence to Non-pharmaceutical interventions regardless of death rate** is incorporated to reflect the population’s will to adhere to Non-pharmaceutical interventions (NPI), as they were known at the time, regardless of a national directive to do so, or at a time when restrictions are being relaxed. This is modelled by assuming that the level of adherence to NPIs in Level 4 (measured by population contact rate) does not increase when restrictions were relaxed to Level 3 and beyond.
d. **Behavioural heterogeneity** acknowledges that some members of society experience different risks and exhibit heterogeneous/ different behavioural patterns, introducing substantial variation in the number of people that different people infect, with highly connected individuals becoming infected earlier in the epidemic and infecting more contacts. This is modelled through adjusting the transmission function (force of infection) to be inflated at the start of the epidemic, but decrease as immunity builds up in the most connected individuals early on.

It is probable that the explanation for the earlier-than-projected plateauing of admissions and deaths at the time in the Western Cape was a combination of these factors, and there was not as yet enough evidence in the international literature or local data for any of these factors (Figure 3). Nonetheless, we ran a number of scenario analyses to see how well these factors would explain the early plateau in the Western Cape, and what the impact of similar phenomena in the three provinces (EC, GP, KZN) with the most progressed epidemics over the next months would be noting that the purpose of the analysis is to demonstrate how each one of these phenomena may be a possible explanation for the observed trends in the Western Cape, rather than attempting to find a best fitting parameter set for each phenomenon (Supplementary table 1).

**Figure 3.**
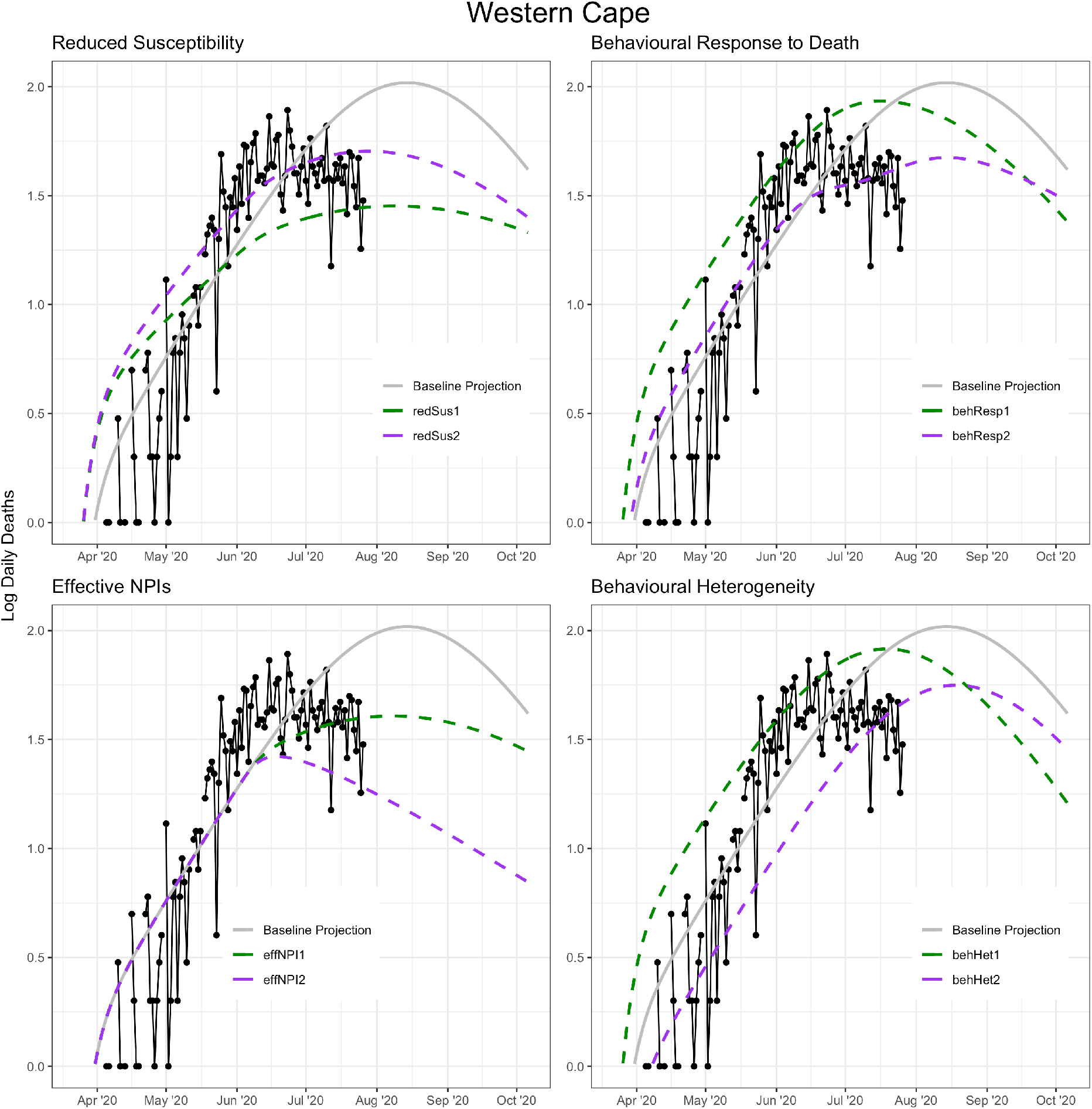
Impact of alternative scenarios on current and projected deaths in the Western Cape

When applying each scenario to the three provinces with the next most advanced epidemics, Eastern Cape, Gauteng and KwaZulu-Natal, we noticed that each in turn led to either later or lower peaking of cases than our original projections, with the exception of the behaviour response to high mortality scenario in which the behavioural response threshold is assumed to be 110 deaths per day, which peaked at roughly the same level but shifted the peak forward slightly in all three provinces (Supplementary Figure 1). Based on these analyses, we chose to include the notion of behavioural heterogeneity as the most plausible explanation as it is a known infectious disease phenomenon that is broadly true of all social contact networks and the model was updated accordingly in the subsequent version of the model.

### NCEM Updated Model Projections

In the original model, we projected the need for hospitalisation based on severity rates, as a means of quantifying the likely burden on the hospital facilities. However with challenges in access to care, and growing estimates of excess deaths, a different approach was considered. Two scenarios were simulated for planning purposes: (1) Use, where admissions to hospital reflect reported use of services with access to care for COVID-19 patients calibrated to 80% of estimated weekly excess deaths, and (2) Need, where *all* those with COVID-19 in need of hospital-level care access and receive it (similar to the original model).

Data-based estimates on length of hospital stay and transitions between stages along the hospital pathways were derived from DATCOV^27^. As 100% of private hospitals but only 53% of public hospitals were participating in DATCOV at the time of the analysis, public sector admissions were underrepresented in this dataset. To adjust for this, we calculated a province-specific inflation factor for the number of general and ICU admissions based on the total number of hospital beds available in both sectors versus the number of beds in hospitals represented in the DATCOV dataset. This inflation factor was applied to the DATCOV admissions data and used for calibration. Both sets of admissions data are presented in the results below, with the inflated data referred to as “adjusted DATCOV data”. This includes the length of hospital stay and the proportion of patients in general vs. ICU wards.

Given the challenges in accurately identifying and recording confirmed COVID-19 deaths, excess deaths provide a more robust measure of mortality. Total projected deaths due to COVID-19 in the model were calibrated to estimates of excess mortality by province from 6 May to 28 August 2020 where 80% of excess deaths were assumed to be due to COVID-19 infection^28^.

Model calibration to hospital admissions and deaths was performed at the provincial level, due to limited district-level data. While all provinces were individually calibrated, provinces with smaller numbers of confirmed cases, hospitalisations and deaths (Free State, Limpopo, Mpumalanga, North West, and Northern Cape) were less easily calibrated. The model code is available under https://sacovid19mc.github.io/, with a full set of provincial model output in the supplementary file. Additionally, district level model output for the Needs scenario has been made available to decision-makers in a dashboard^29^.

As testing guidelines and practices changed between provinces and over time as best practices on testing emerged with the limited supply of test kits and testing backlogs, cumulative detected cases were estimated under two scenarios i) moderate testing coverage as implemented in May and June, and ii) a more limited testing coverage policy implemented from mid-June that prioritised testing in hospitalised cases and in healthcare workers. Figure 5 summarises the projected cumulative detected cases at the national level assuming the current testing policy (blue) and a limited policy of detecting only hospitalised cases (orange). Importantly, a change in the testing policy only affects the number of detected cases, not any of the other projections. Table 1 gives an overview of the projections at select dates.

**Figure 4.**
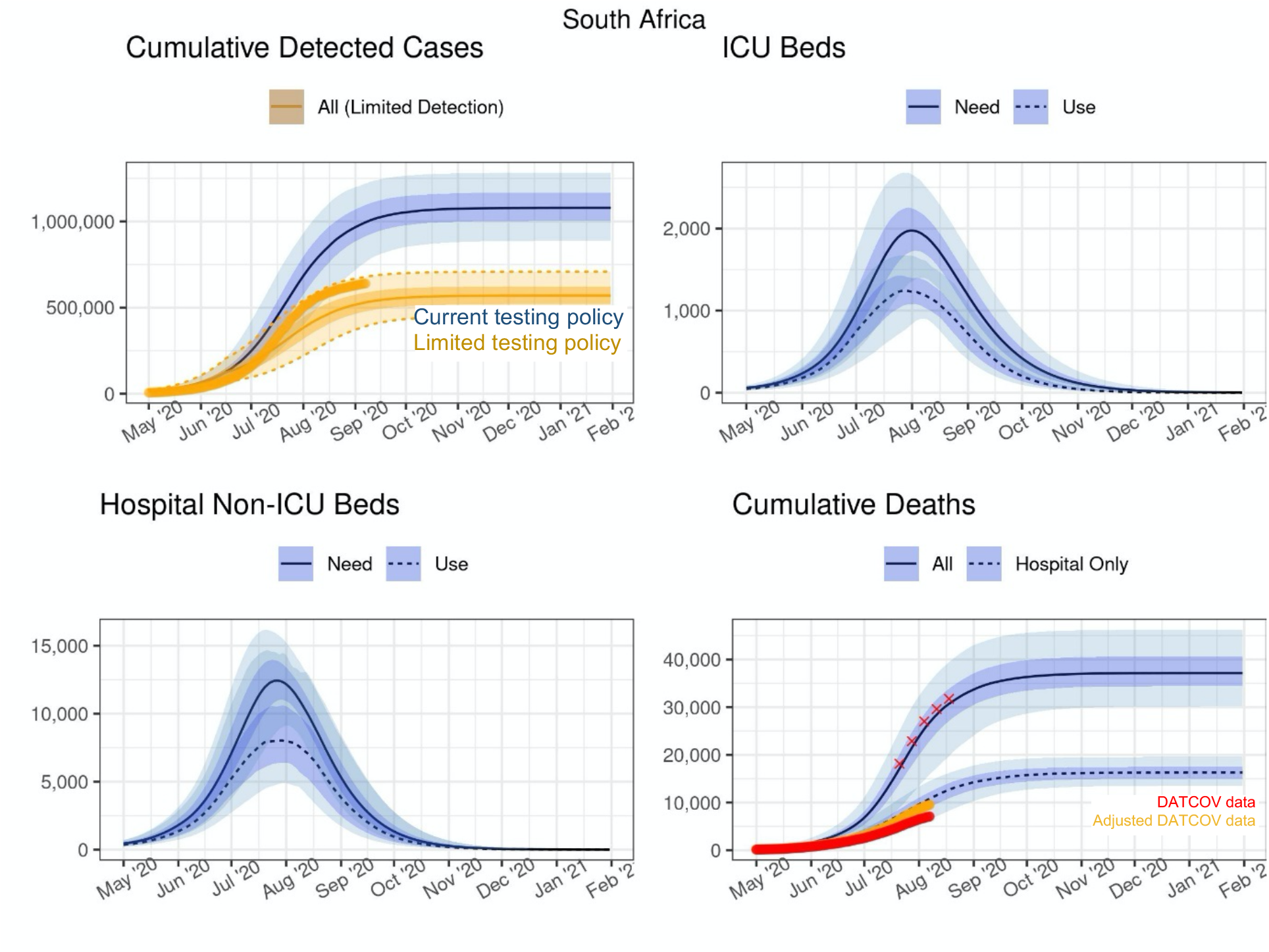
Projected cases and inpatient bed need and use at the national level. The red crosses in the bottom right-hand panel represents 80% of the excess deaths found in the SAMRC analysis

**Figure 5.**
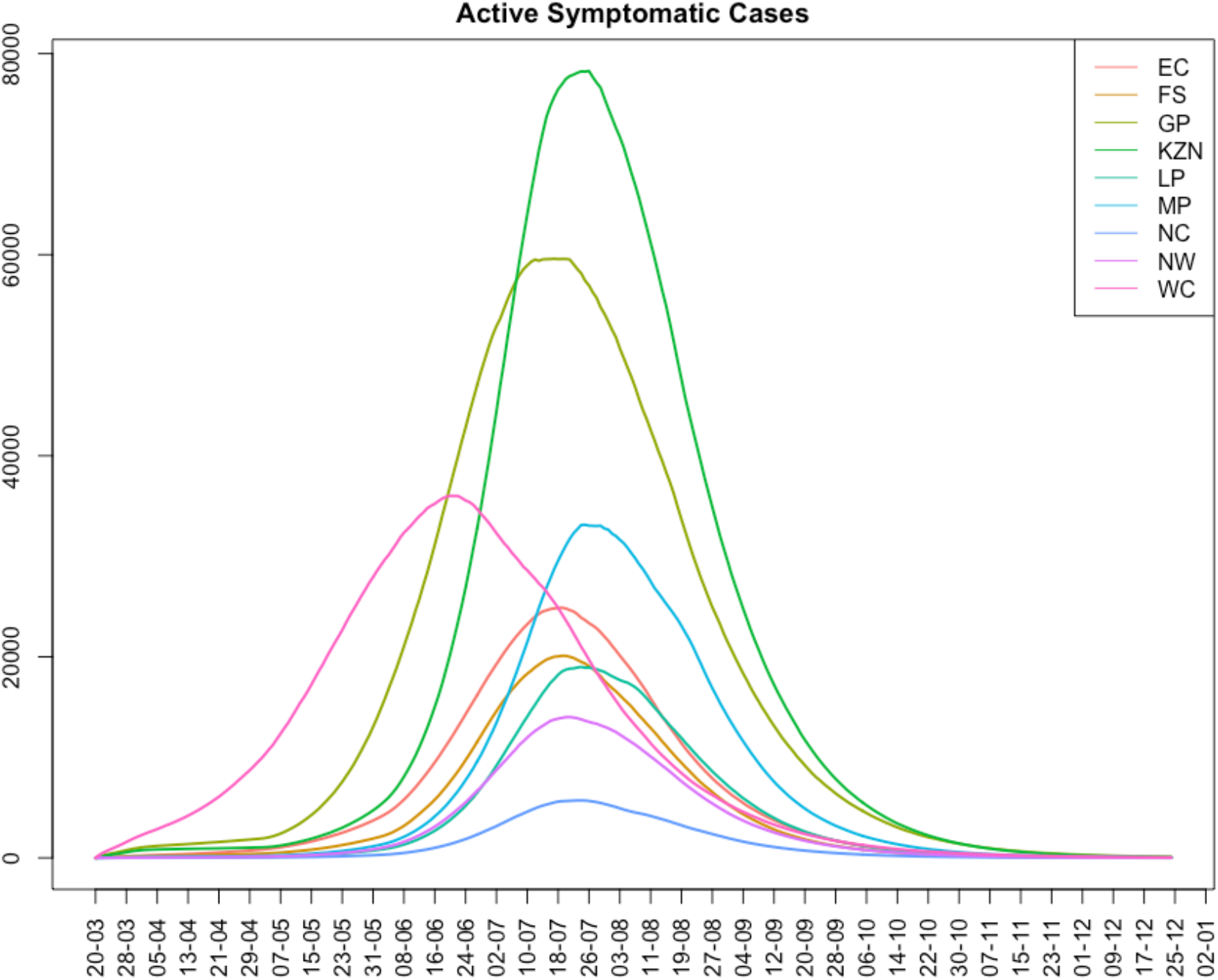
Estimated daily active symptomatic cases by province from week beginning 20 March 2020 to 25 December 2020

**Table 1.** Projections of national cases, deaths and admissions needed at select dates

| Table for SA<br>2020-09-01 to 2021-02-02 |  |  |  |  |  |
| --- | --- | --- | --- | --- | --- |
| Date | Cumulative Incidence |  | Active Cases |  | Cumulative Detected Cases |
|  | Total | Symptomatic | All | Symptomatic | Limited Detection |
| 2020-09-01 | 15,201,000<br>(13,138,000 - 17,023,000) | 3,697,800<br>(2,835,700 - 4,521,500) | 462,600<br>(267,000 - 817,800) | 85,260<br>(46,810 - 151,500) | 517,000<br>(372,000 - 668,300) |
| 2020-10-01 | 15,840,000<br>(14,112,000 - 17,488,000) | 3,914,700<br>(3,172,100 - 4,688,100) | 102,300<br>(54,020 - 238,000) | 19,320<br>(9,730 - 47,560) | 557,800<br>(433,500 - 700,400) |
| 2020-11-01 | 15,977,000<br>(14,415,000 - 17,581,000) | 3,961,300<br>(3,246,400 - 4,720,300) | 20,840<br>(10,210 - 52,710) | 3,950<br>(1,840 - 10,690) | 567,500<br>(447,800 - 707,100) |
| 2020-12-01 | 16,004,000<br>(14,480,000 - 17,596,000) | 3,970,400<br>(3,260,300 - 4,727,800) | 4,510<br>(2,050 - 11,990) | 860<br>(370 - 2,500) | 569,300<br>(450,400 - 708,600) |
| 2021-01-01 | 16,010,000<br>(14,499,000 - 17,599,000) | 3,972,000<br>(3,263,400 - 4,729,700) | 950<br>(390 - 2,700) | 180<br>(70 - 570) | 569,800<br>(451,000 - 708,900) |
| 2021-02-01 | 16,011,000<br>(14,501,000 - 17,600,000) | 3,972,400<br>(3,264,000 - 4,730,000) | 210<br>(70 - 650) | 40<br>(10 - 140) | 569,900<br>(451,200 - 709,000) |

| Date | Cumulative Admissions |  | Hospital Beds in Use |  | Cumulative Deaths |  |
| --- | --- | --- | --- | --- | --- | --- |
|  | General | ICU | Non-ICU | ICU | Hospital | All |
| 2020-09-01 | 97,500<br>(73,290 - 121,400) | 11,940<br>(9,010 - 15,060) | 3,830<br>(1,720 - 8,410) | 720<br>(400 - 1,090) | 14,250<br>(10,300 - 17,760) | 33,710<br>(24,020 - 43,010) |
| 2020-10-01 | 105,700<br>(85,730 - 127,400) | 12,950<br>(10,550 - 15,860) | 950<br>(360 - 2,810) | 200<br>(90 - 430) | 15,740<br>(12,680 - 19,210) | 36,330<br>(28,640 - 45,600) |
| 2020-11-01 | 107,800<br>(88,740 - 128,900) | 13,190<br>(10,900 - 16,110) | 190<br>(70 - 680) | 40<br>(10 - 130) | 16,170<br>(13,320 - 19,510) | 37,000<br>(29,850 - 46,140) |
| 2020-12-01 | 108,200<br>(89,310 - 129,300) | 13,240<br>(10,970 - 16,150) | 40<br>(10 - 160) | <10<br>(<10 - 40) | 16,260<br>(13,460 - 19,570) | 37,140<br>(30,120 - 46,230) |
| 2021-01-01 | 108,300<br>(89,420 - 129,400) | 13,250<br>(10,980 - 16,160) | <10<br>(<10 - 40) | <10<br>(<10 - <10) | 16,290<br>(13,490 - 19,580) | 37,160<br>(30,170 - 46,260) |
| 2021-02-01 | 108,300<br>(89,450 - 129,400) | 13,260<br>(10,980 - 16,160) | <10<br>(<10 - <10) | <10<br>(<10 - <10) | 16,300<br>(13,500 - 19,580) | 37,170<br>(30,180 - 46,260) |

Under the moderate testing scenario, cumulative detected cases were projected to continue to grow until 1.2 million in early November 2020, and only marginally so thereafter, whereas only approximately 567,500 cases (447,800-707,100) were estimated to be detected under limited testing. By September, the reported number of detected cases had already surpassed the median of the limited testing scenario and over 725,000 detections by November 2020.

The COVID-19 epidemic was estimated to have peaked nationally in mid-July with approximately 16,000,000 infections by December 2020, representing 26.8% (24.3%, 29.5%) of the population. Total deaths were estimated to continue to increase until early November when the cumulative number of all deaths would reach 37,000 (of which 16,000 will have been in hospital); thereafter the growth rate was estimated to be very low (Figure 5).

For the estimation of hospital bed requirements, scenarios of both the estimated *need* and the actual *use* of ICU and non-ICU beds are depicted. The peak number of general hospital (i.e., non-ICU) beds *in use* was estimated to be reached in early-August, at around 8,000 beds (when around 12,500 beds were estimated to have been *needed*). The peak number of ICU beds in use was estimated to be reached around the same time, with around 1,100 beds (when more than 2,000 beds would have been needed) (Figure 4).

At a provincial level, considerable variation was observed and projected in the timing and height of peak infection between the provinces (Figure 6). This allowed the strain on healthcare resources to be spread out, potentially allowing for more healthcare capacity based on how well resources such as beds, oxygen, test kits and reagents and staff could be shifted between provinces (and within provinces, patients needing hospitalisation could be moved between under-and better resourced districts).

**Figure 6.**
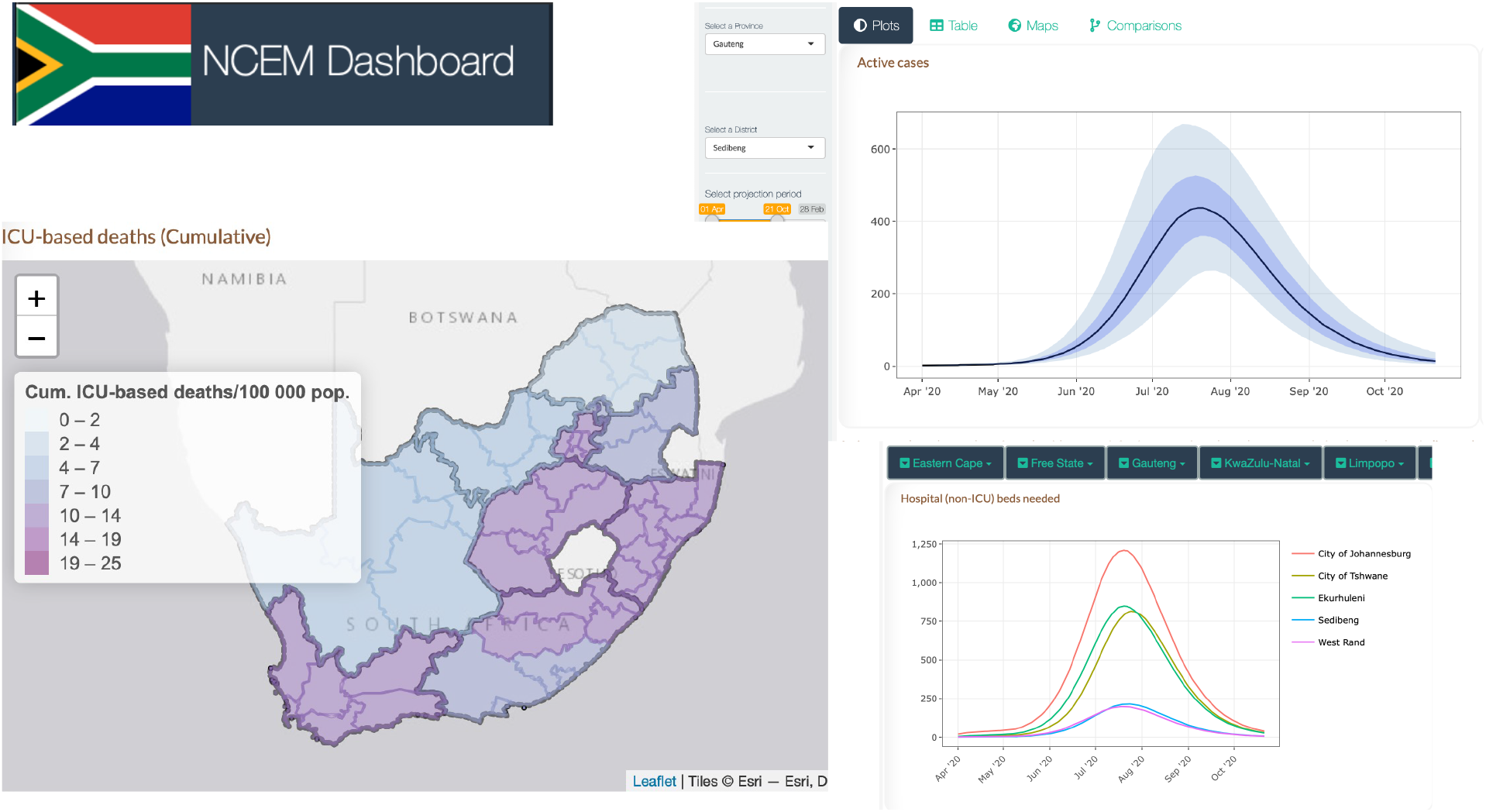
NCEM Dashboard Sample Output: Interactive dashboard for district level projections of cases, admissions and deaths as at 11 September 2020^29^

For all provinces, projections of deaths and cases requiring hospitalisation are presented in Supplementary figures 2-10 and Tables 3-11. Across provinces, estimates of all COVID-19 related deaths were projected to be almost double those of the reported COVID-19 related deaths occurring in hospital (which the DATCOV hospital surveillance dataset aims to capture). For example, in the Eastern Cape Province (Supplementary Figure 2), adjusting for hospital representativeness in the DATCOV database, hospital-based deaths were projected to reach 2500 by October 2020, while total deaths were estimated at more than 7,500 at the same time-assuming 80% of excess deaths were due to COVID-19. This differential is additionally reflected in the difference in the projected need for and use of hospital services such that use of hospital beds was estimated to peak at 100 and 750 median ICU and non-ICU beds respectively whereas if all patients who needed care received it, the estimated need for hospital beds would be 200 and 1500 median ICU and non-ICU beds respectively.

The NCEM dashboard provides the user with the ability to interact with projections of cases, hospitalisations and deaths for the first wave of the epidemic across the 52 districts in South Africa^29^. Figure 6 shows a sample of outputs such as maps, comparison line plots, tables and reports at the district level, where the district-level waves show additional variation within provinces.

## Discussion

Mathematical modelling plays a role in contributing to both improving the understanding of the disease, and improving planning in order to make better decisions and reduce disease impact. The model presented here has been developed using data that is subject to a high degree of uncertainty. As all models are simplifications of reality that are designed to describe and predict system behaviour and are a product of the assumptions and data with which they are developed, the model used to support planning in South Africa was refined regularly throughout the South African COVID-19 epidemic^14^.

COVID-19 modelling groups around the world had various levels of impact on a country or countries’ policy choices. In South Africa, given our consortium’s early mandate from the National Department of Health, the government department tasked with coordinating the country’s COVID-19 response, we were able to work very closely with decision makers and planners at all levels, from two of the three Ministerial Advisory Committees on COVID-19 advising the Minister of Health to National Treasury to the individual teams tasked with planning facility readiness and, from 2021 on, the vaccine programme. Examples of uses of model projections included the estimation of drug quantities for in-patient and out-patient care at facilities; the quantification of additional mortuary and burial spaces; the cost of ventilation equipment for all hospitals; estimating the required supply of oxygen, planning of district facility space, scenario planning of interventions, and supporting the analysis of the macroeconomic impact of the epidemic under different scenarios by the Reserve Bank.

Publicly-accessible dashboards visualising the main results of NCEM outputs for the first wave^29^ and our resurgence monitoring metrics and 2-week forecasts^30^ additionally allowed us to make central model results available to the public. Continuously updating model assumptions, parameters and projections over the course of the epidemic and communicating changes through an established communication pipeline to planners across the different levels of government, lead to increased credibility and reliance on model outputs as a signal for planning. This was further demonstrated across the next three waves of infections where model projections were routinely requested to assist with planning.

The NCEM attempted to incorporate a high level of local data to contextualise the model appropriately to address South Africa’s population and health system characteristics. The data however, were not always representative of the full population or complete. For example, in many countries, reported SARS-CoV-2 related deaths were only a fraction of the total excess deaths seen during the last two years. South Africa is one of the few African countries with a vital statistics reporting system that allows the estimation of excess deaths. This put us in the unusual situation for a LMIC to be able to estimate SARS CoV-2 mortality in more detail. COVID-19-specific mortality reports in South Africa were largely limited to within-hospital deaths, with public sector hospitals dependent on manual capturing of confirmed COVID-19 deaths. Under-reporting of COVID-19 mortality arose from individuals not having been tested for SARS-CoV-2 before dying, especially when dying outside of hospitals or care facilities, and incomplete reporting of in-hospital deaths.

Pressure on data-capturing systems during the peaks of waves additionally likely resulted in under-reporting of both confirmed COVID-19 admissions and deaths. Furthermore, access to health services in South Africa is variable. Less than 60% of poor households have available, affordable and acceptable access to health services, with rural communities having lower access than urban communities^31^. Less than half of influenza deaths are medically attended each year^32^.

Our projections at the district level did not capture low-level clustering of cases. The population level model made simplifying assumptions regarding how contacts between infectious and uninfected people occur through grouping individuals at the district level. It does not also capture the effects of specific events on local transmission. Though contact rates in each district were adjusted uniformly based on national policy changes, stochastic simulation allowed for variation at each time step for each state transition. Additionally, the 6-month projections during the first wave of infections failed to take into account the possibility of new variants at the time (as variants of SARS-CoV-2 had not yet been detected), which ultimately have a substantial impact on the trajectory of the epidemic a few months after the end of the first wave.

The role of population behaviour in determining the trajectory and scale of the epidemic is more influential in COVID-19 due to the absence of a cure or vaccine at the time of developing the model. It is not possible to precisely capture human emotion and behaviour varied on a fine spatial scale in an equation, least of all in an unprecedented situation such as the current pandemic. Accounting for these unknowns requires that models are run stochastically and estimates are presented with uncertainty bands reflective of variation in the parameters driving the model and the model process itself.

The NCEM is currently being adapted and implemented to assess the impact of vaccine and infection-derived immunity on the likely severity of future infection. However, the future of the spread of SARS-CoV-2 and the impact of COVID-19 on health and health resources depends on many unknowns, including the duration of infection- and vaccine-derived immunity. Depending on the nature of immunity from either past infection or vaccination, the future of SARS-CoV-2 could become regular annual epidemics caused by novel variants, seasonal epidemics, epidemics occurring every few years or even sporadic, unpredictable epidemics. It is therefore important to continue to monitor the epidemic and remain vigilant to detect localised outbreaks as and when they occur.

## Conclusion

The NCEM has been developed as a tool to project cases and deaths due to COVID-19. Its features allow for transmission to be modelled at a fine spatial scale and population behaviour to be captured through mobility, contact patterns and adherence to non-pharmaceutical and pharmaceutical interventions. The model presented in this manuscript has demonstrated the need to adapt models rapidly to changing local data and knowledge in order to support government planning needs. The model has since been adapted to include variants and vaccines and calibrated to additional waves of COVID-19.

The pandemic presented the opportunity for infectious disease modellers around the world to develop tools with speed. Pandemic preparedness endeavours need to incorporate provisions for increasing modelling capacity and training of future modellers, particularly in LMIC. These should be prioritised by local governments and donors alike.

## Data Availability

All model code available under https://sacovid19mc.github.io/

## Acknowledgements

The authors acknowledge the support of the National Department of Health and all planners, stakeholders and decision-makers who made use of these data systems and analyses to respond to the COVID-19 epidemic in South Africa. A large number of colleagues within the National Institute of Communicable Diseases, the Ministerial Advisory Committee on COVID-19, the Clinton Health Access Initiative, provincial and national Departments of Health, and National Treasury alerted us to policy-relevant questions, assisted the development of our models by providing data, discussing details of model structure, and reviewing assumptions and results. We are especially grateful to decision makers and planners at all levels of the health system who continuously used our outputs in the management of COVID-19.

## Ethics statement

Ethical clearance for the modelling study with access to third party data was granted by the University of Cape Town Faculty of Science Research Ethics Committee (FSREC_003_2001) and Stellenbosch University (project ID 19330, ethics reference no.

N20/11/074_RECIP_WITS_M160667_COVID-19).

## Supplementary material

Additional outputs : Supplementary file.doc

Model code: https://sacovid19mc.github.io/

