## Supplementary File for "The National COVID-19 Epi Model (NCEM): Estimating cases, admissions and deaths in South Africa"

### Model Performance and Scenario analysis

We performed a number of scenario analyses to interrogate the impact that four distinct factors could have had in explaining the difference between the NCEM projections from June and reported case and death data in the Western Cape.

These were:

a) A **lower than assumed population attack rate**, possibly due to different levels of susceptibility in different population groups (including children) or the presence of existing T-cell derived-immunity after prior exposure to other coronaviruses. This is modelled by allowing a proportion of individuals to be immune throughout the course of the epidemic.

b) **Behaviour change** **in response to an increased local death rate**. This scenario takes into account a potential impact of public awareness of the increasing deaths and the looming threat of overwhelmed healthcare facilities in the Western Cape, which, combined with communication campaigns, may have resulted in better adherence to non-pharmaceutical interventions (NPIs) (e.g. masks, hand washing and physical distancing) and in those most at risk for severe COVID-19 disease taking additional precautions to isolate themselves. This is modelled by allowing the population in each district to reduce interactions when district death rates are high and increase interactions when death rates are low.

c) **Better adherence to Non-pharmaceutical interventions regardless of death rate** is incorporated to reflect the population’s will to adhere to Non-pharmaceutical interventions **(**NPI), as they were known at the time, regardless of a national directive to do so, or at a time when restrictions are being relaxed. This is modelled by assuming that the level of adherence to NPIs in Level 4 (measured by population contact rate) does not increase when restrictions were relaxed to Level 3 and beyond.

d) **Behavioural heterogeneity** acknowledges that some members of society experience different risks and exhibit heterogeneous/ different behavioural patterns, introducing substantial variation in the number of people that different people infect, with highly connected individuals becoming infected earlier in the epidemic and infecting more contacts. This is modelled through adjusting the transmission function (force of infection) to be inflated at the start of the epidemic, but decrease as immunity builds up in the most connected individuals early on.

Table 1 summarises how we implemented each of these scenarios by investigating the impact of different model parameters.

Table 1: Scenario parameters

| **Scenario** | **Description** |
| --- | --- |
| ***Reduced susceptibility*** | |
| redSus1 | 12.5% of the population assumed to be completely immune to infection. Additional curvature achieved by assuming a further 20% reduction in contacts from Level 3 restrictions. (Asymptomatic proportion: 0.75) |
| redSus2 | 6% of the population assumed to be completely immune to infection. (Asymptomatic proportion: 0.75)  Note that different combinations of asymptomatic proportion and immune proportion can yield similar results. |
| ***Behaviour response to high mortality*** | |
| behResp1 | Half-saturation point / response threshold is assumed to be 110 deaths per day |
| behResp2 | Half-saturation point / response threshold is assumed to be 30 deaths per day with a reduced seed |
| ***Better adherence to NPIs*** | |
| effNPI1 | Average contacts during level 4 decreased to 80% during level 3 and beyond |
| effNP!2 | Average contacts during level 4 decreased to 65% during level 3 and beyond |
| ***Behavioural heterogeneity*** | |
| behHet1 | Concavity parameter k = 0.3, with increased seed |
| behHet2 | Concavity parameter k=1, with reduced seed |

Figure 1 Impact of alternative scenarios on current and projected deaths in the Eastern Cape, Gauteng and KwaZulu-Natal

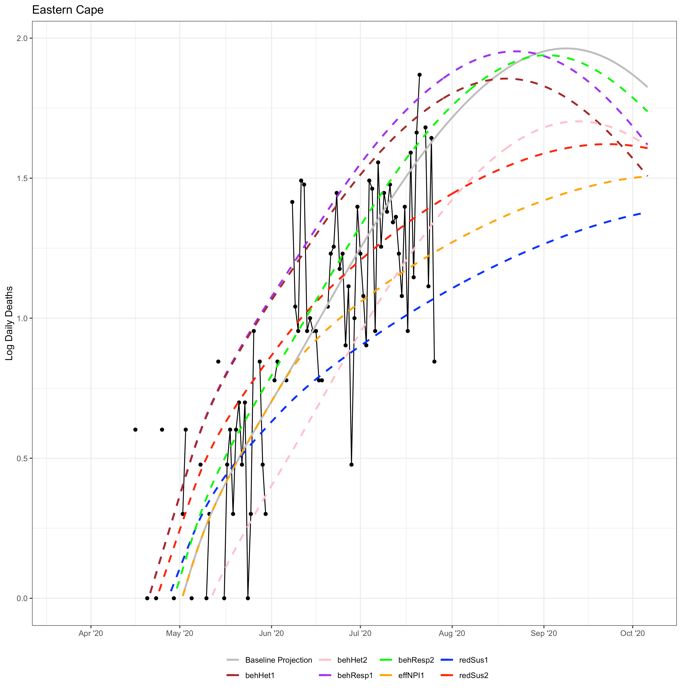

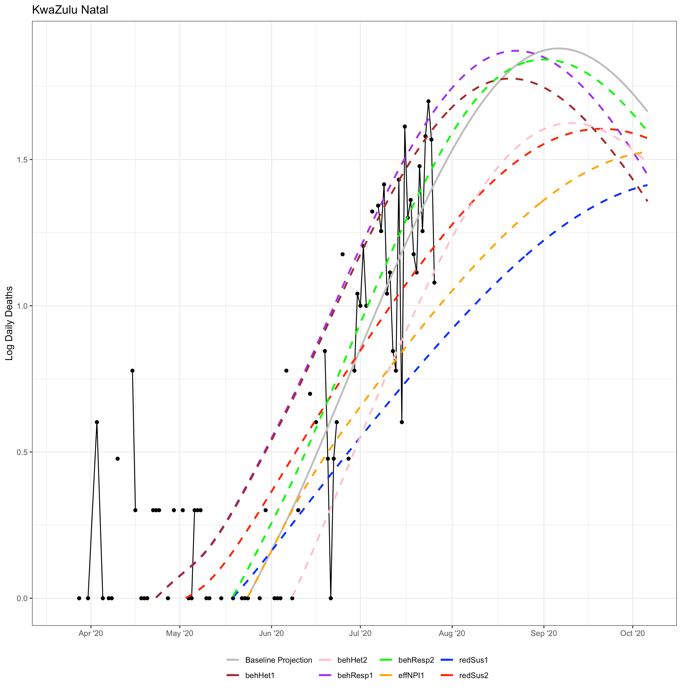

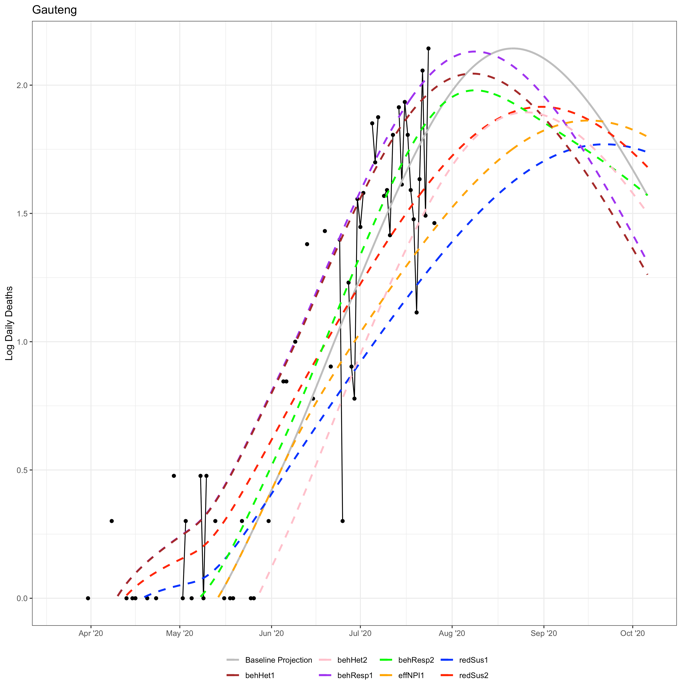

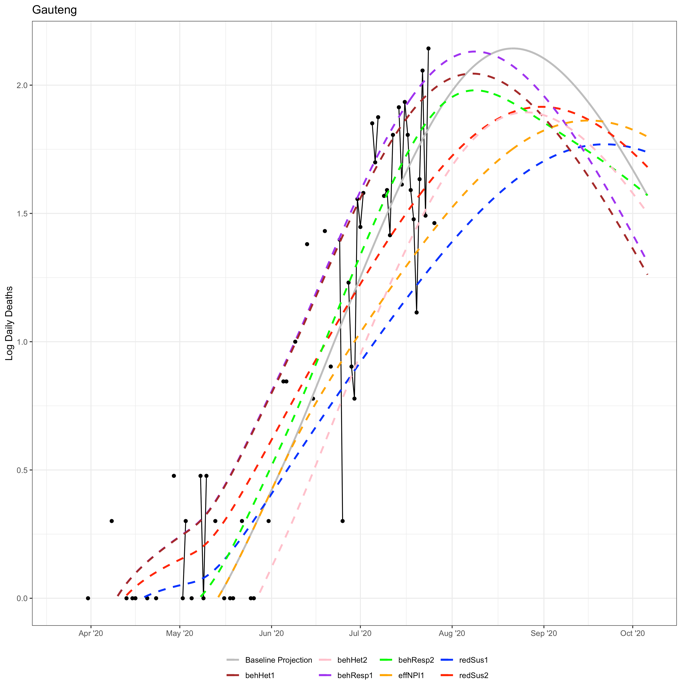

### NCEM v4.0 (District Model): Supplementary information

This document provides a technical overview of the National COVID-19 Epi Model (NCEM) provincial model. The model described in this document is the district-level NCEM model, created by the [South African COVID-19 Modelling Consortium](https://saCOVID19mc.github.io). There is also a separate document, the [NCEM Provincial Model Code Guide](https://sacovid19mc.github.io/ncemProvincialCodeGuide.pdf), that gives an overview of the structure of the model code for the Provincial Model; the district model code has a similar structure. If there are any queries regarding the model or the code, please contact us on:.

#### Model equations

The model describes the temporal evolution of the following state variables:

| Variable | Definition |
| --- | --- |
| $S$ | number of susceptible individuals |
| $E$ | number of exposed but not yet infectious individuals |
| $I_{A}$ | number of asymptomatic individuals (infectious) |
| $I_{P}$ | number of presymptomatic individuals (infectious) |
| $I_{M}$ | number of mildly and moderately ill individuals (infectious) |
| $I_{ST}$ | number of individuals who are or will become severely ill and will access treatment but are not yet hospitalised (infectious) |
| $I_{S\bar{T}}$ | number of individuals who are or will become severely ill but will not access treatment (infectious) |
| $H_{1}$ | number of severely ill individuals who are hospitalized in the general (non-ICU) ward |
| $H_{2}$ | number of individuals who are work will be come critically ill currently in the general (non-ICU) ward |
| $C_{V1}$ | number of individuals who are critically ill, will eventually die, and are currently in the ICU (ventilator resourced) |
| $C_{V2}$ | number of individuals who are critically ill, will eventually recover, and are currently in the ICU (ventilator resourced) |
| $C_{\bar{V}1}$ | number of individuals who are critically ill, will eventually die, and are currently in the ICU (not ventilated) |
| $C_{\bar{V}2}$ | number of individuals who are critically ill, will eventually recover, and are currently in the ICU (not ventilated) |
| $H_{3}$ | number of individuals who have been critically ill, will recover, and have been discharged from the ICU but remain in hospital for step-down care |
| $W_{H}$ | number of severely ill individuals who have sought hospitalization but could not be accommodated |
| $W_{V}$ | number of critically ill individuals who require a ventilator-resourced ICU bed but could not be accommodated |
| $W_{\bar{V}}$ | number of critically ill individuals who require a non-ventilator-resourced ICU bed but could not be accommodated |
| $R$ | number of individuals who are no longer infectious / recoverd and/or discharged |
| $D$ | number of individuals who have died |
| $I_{M_{d}}$ | cumulative number of *confirmed* mild / moderate infections |
| $I_{S_{d}}$ | cumulative number of *confirmed* severe and critical infections |
| $N$ | total number of individuals in the population ($S+E+I_{A}+I_{P}+I_{M}+I_{ST}+I_{S\bar{T}}+H_{1}+H_{2}+C_{V1}+C_{V2}+C_{\bar{V}1}+C_{\bar{V}2}+H_{3}+R+W_{S}+W_{V}+W_{\bar{V}}$) |
| $X$ | dummy variable representing mild and moderate cases who will be tested before they are tested |
| $Y$ | dummy variable representing severe and critical cases who will be tested before they are tested |

The following equations describe the dynamics of transmission and disease progression within each district, $x$:

$$\frac{dS_{x}}{dt}=-\Phi_{x}S_{x}$$

$$\frac{dE_{x}}{dt}=\Phi_{x}S_{x}-\gamma_{1}E_{x}$$

$$\frac{dI_{A_{x}}}{dt}=p_{a}\gamma_{1}E_{x}-r_{1}I_{A_{x}}$$

$$\frac{dI_{P_{x}}}{dt}=(1-p_{a})\gamma_{1}E_{x}-\gamma_{2}I_{P_{x}}$$

$$\frac{dI_{M_{x}}}{dt}=p_{m_{x}}\gamma_{2}I_{P_{x}}-r_{2}I_{M_{x}}$$

$$\frac{dI_{{ST}_{x}}}{dt}=(1-p_{m_{x}})p_{t_{x}}\gamma_{2}I_{P_{x}}-\tau_{s}I_{{ST}_{x}}$$

$$\frac{dI_{{S\bar{T}}_{x}}}{dt}=(1-p_{m_{x}})(1-p_{t_{x}})\gamma_{2}I_{P_{x}}-r_{8}I_{{S\bar{T}}_{x}}$$

$$\frac{dH_{1_{x}}}{dt}=\alpha_{1_{x}}\left( 1-\frac{p_{c_{x}}}{1-p_{m_{x}}} \right)\tau_{s}I_{{ST}_{x}}-r_{3}H_{1_{x}}$$

$$\frac{dH_{2_{x}}}{dt}=\alpha_{1_{x}}\frac{p_{c_{x}}}{1-p_{m_{x}}}\tau_{s}I_{{ST}_{x}}-\tau_{p}H_{2_{x}}$$

$$\frac{dC_{{V1}_{x}}}{dt}=\alpha_{2_{x}}p_{v_{x}}d_{{cv}_{x}}\tau_{p}H_{2_{x}}-\mu_{v}C_{{V1}_{x}}$$

$$\frac{dC_{{V2}_{x}}}{dt}=\alpha_{2_{x}}p_{v_{x}}(1-d_{{cv}_{x}})\tau_{p}H_{2_{x}}-r_{9}C_{{V2}_{x}}$$

$$\frac{dC_{\bar{V1}_{x}}}{dt}=\alpha_{2_{x}}(1-p_{v_{x}})d_{{c\bar{v}}_{x}}\tau_{p}H_{2_{x}}-\mu_{\bar{v}}C_{\bar{V}1}$$

$$\frac{dC_{{\bar{V}2}_{x}}}{dt}=\alpha_{2_{x}}(1-p_{v_{x}})(1-d_{{c\bar{v}}_{x}})\tau_{p}H_{2_{x}}-r_{10}C_{{\bar{V}2}_{x}}$$

$$\frac{dH_{3_{x}}}{dt}=r_{9}C_{{V2}_{x}}+r_{10}C_{{\bar{V}2}_{x}}-r_{5}H_{3_{x}}$$

$$\frac{dR_{x}}{dt}=r_{1}I_{A_{x}}+r_{2}I_{M_{x}}+(1-d_{s_{x}})r_{3}H_{1_{x}}+r_{5}H_{3_{x}}+\left( 1-\frac{p_{c_{x}}d_{C\bar{T}}}{1-p_{m_{x}}}-\frac{p_{s_{x}}d_{S\bar{T}}}{1-p_{m_{x}}} \right)r_{8}(W_{H_{x}}+I_{{S\bar{T}}_{x}})$$

$$\frac{dD_{x}}{dt}=d_{s_{x}}r_{3}H_{1_{x}}+\mu_{V}C_{{V1}_{x}}+\mu_{\bar{V}}C_{{\bar{V}1}_{x}}+\left( \frac{p_{c_{x}}d_{C\bar{T}}}{1-p_{m_{x}}}-\frac{p_{s_{x}}d_{S\bar{T}}}{1-p_{m_{x}}} \right)r_{8}(W_{H_{x}}+I_{{S\bar{T}}_{x}})+r_{9}W_{V_{x}}+r_{10}W_{\bar{V}_{x}}$$

$$\frac{dW_{H_{x}}}{dt}=(1-\alpha_{1})\tau_{s}I_{ST}-r_{8}W_{H_{x}}$$

$$\frac{dW_{V_{x}}}{dt}=(1-\alpha_{2})p_{v_{x}}\tau_{p}H_{2}-r_{9}W_{V_{x}}$$

$$\frac{dW_{\bar{V}_{x}}}{dt}=(1-\alpha_{2})(1-p_{v_{x}})\tau_{p}H_{2}-r_{10}W_{\bar{V}_{x}}$$

$$\frac{dX_{x}}{dt}=d_{m}p_{m_{x}}\gamma_{2}I_{P_{x}}-\Delta_{m}X_{x}$$

$$\frac{dY_{x}}{dt}=\alpha_{1}d_{s}\tau_{s}I_{{ST}_{x}}-\Delta_{s}Y_{x}$$

$$\frac{dI_{M_{d}}}{dt}=\Delta_{m}X_{x}$$

$$\frac{dI_{S_{d}}}{dt}=\Delta_{s}Y_{x}$$

where the force of infection, $\Phi_{x}$, is defined as

$$\Phi_{x}=\frac{\beta_{x}\delta_{x,t}\left( \zeta I_{A_{x}}+\zeta I_{P_{x}}+I_{M_{x}}+I_{{ST}_{x}}+I_{{S\bar{T}}_{x}}+W_{H_{x}} \right)}{N_{x}}$$

and $p_{c_{x}}=1-p_{m_{x}}-p_{s_{x}}$.

#### Key parameter values

Tables 2 and 3 below show the values of key parameters used to inform the model. Parameter values have been selected for use by an expert panel of clinicians on the SA COVID-19 Modelling Consortium and updated with inputs from recent South African data where indicated. Parameter values that are provided as ranges only differ by province.

Table 2 Results of NICD analysis of estimated national and provincial reproductive numbers [1,3]*

| **Restriction level** | **National** | **Eastern Cape** | **Gauteng** | **KwaZulu Natal** | **Western Cape** |
| --- | --- | --- | --- | --- | --- |
| None (R_0_) | 2.5 (2, 3) | 2.5 (2, 3) | 2.5 (2, 3) | 2.5 (2, 3) | 2.5 (2, 3) |
| Level 5 R_t_^2^ | 1.3  (1.0, 1.6) | 1.4  (1.1, 1.7) | 1.2  (1.0, 1.4) | 1.1  (0.9, 1.43 | 1.5  (1.2, 1.8) |
| Level 4 R_t_: NICD R_t_ estimates calibrated to fit hospital-based provincial deaths^2^ | 1.6  (1.3, 1.9) | 1.6  (1.2, 1.8) | 1.8  (1.4, 2.2) | 1.6  (1.3, 1.9) | 1.6  (1.3, 1.9) |
|  | **Other Provinces** | **Eastern Cape** | **Gauteng** | **KwaZulu Natal** | **Western Cape** |
| Level 3 (1–30 June): increase in contacts (relative to previous period) estimated from a decrease in residential mobility^3^ | (21.1%, 26.0%) | 21.0%  (16.8, 25.2) | 4.3%  (3.4, 5.2) | 20.5%  (16.4, 24.6) | 8.1%  (6.5, 9.7) |
| Level 3 (1 July – 17 August): increase in contacts (relative to previous period) estimated from a decrease in residential mobility^3^ | (0.5%, 2.5%) | 3.0%  (2.4, 3.6) | 1.8%  (1.4, 2.2) | 0.8%  (0.6, 1.0) | 6.6%  (5.3, 7.9) |
| Level 2 (18 August ->): increase in contacts (relative to previous period) estimated from a decrease in residential mobility^3^ | (3.3%, 5.8%) | 5.4%  (4.3, 6.5) | 1.7%  (1.4, 2.0) | 4.8%  (3.8, 5.8) | 3.1%  (2.5, 3.7) |

* We utilised national estimates where provincial data was too sparse. R_0_, and R_t_ for Level 5 and Level 4 from symptom onset date adjusted for testing volumes

Table 3. Key model parameters

|  | Parameter | Value (range) | Sources |
| --- | --- | --- | --- |
| Infection severity | Proportion of cases that are asymptomatic | 75% (70% - 80%) | [9-12] |
|  | Relative infectiousness of asymptomatic cases | 80% (77.5%, 82.5%) | [13-15]  Estimated through calibration to admissions and fatalities count data (DATCOV) [4] |
|  | Mild to moderate cases among the symptomatic | (94.55% - 97.13%) | Estimated through calibration to admissions and fatalities count data (DATCOV) [4] |
|  | Severe cases among the symptomatic | (2.58% - 5.00%) |  |
|  | Critical cases among the symptomatic | (0.18% - 0.55%) |  |
|  | Fatal cases among the admitted (general) | (6.82% - 20.28%) | Estimated from NICD COVID-19 Hospital Sentinel Surveillance database (DATCOV) [4] & Western Cape Line List Data (SPV) [16] |
|  | Fatal cases among the admitted (ICU ventilated) | (43.01% - 85.03%) |  |
|  | Fatal cases among the admitted (ICU non-ventilated) | (22.73% - 43.35%) |  |
|  | Proportion of cases in ICU requiring ventilation | (19.44% - 51.47%) |  |
|  | Fatal cases among the critically infected requiring ventilation, *in the absence of appropriate care* | 100% | Expert opinion of clinicians convened by the National COVID-19 Modelling Consortium |
|  | Fatal cases among the critically infected not requiring ventilation, *in the absence of appropriate care* | Unchanged: Fatal cases among the admitted (ICU non-ventilated) |  |
|  | Fatal cases among the critically infected requiring oxygen, *in the absence of appropriate care* | 100% |  |
|  | Fatal cases among the severely infected requiring oxygen, *in the absence of appropriate care* | 90% |  |
|  | Probability of seeking hospital-level care for severely and critically ill | (50.00% - 97.00%) | Estimated through calibration to 80% of excess mortality [5] |
| Timeframes & treatment durations | Time from infection to onset of infectiousness | 2 days (1.0 - 3.0) | [8, 17-26]  with input from the National COVID-19 Modelling Consortium |
|  | Time from onset of infectiousness to onset of symptoms | 4 days (3.0 - 5.0) |  |
|  | Duration of infectiousness from onset of symptoms | 5 days (4.0 - 6.0) | [26, 27] |
|  | Time from onset of symptoms to testing | 4 days (3.0 - 5.0) | [17,18, 28-32] |
|  | Time from onset of symptoms to hospitalisation | 5 days (4.0 - 6.0) |  |
|  | Time in non-ICU (never ICU) to death/recovery | 8 days (4.0 - 12·0) | Lengths of stay: values and ranges sourced from NICD COVID-19 Hospital Sentinel Surveillance database (DATCOV) [4] |
|  | Time in non-ICU for those destined for ICU | 0 days (0.0 - 2.0) |  |
|  | Time in ICU for those ventilated and destined to die | 14 days (7.0 - 27.0) |  |
|  | Time in ICU for those never ventilated and destined to die | 11 days (7.0 - 18.0) |  |
|  | Time in ICU for those ventilated and recovered | 19 days (15.0 - 37.0) |  |
|  | Time in ICU for those never ventilated and recovered | 5 days (1.0 - 10.0) |  |
|  | Time in non-ICUs for those who were in ICU and recovered | 0 days (0.0 - 6.0) |  |

^*^ A full list of parameters are available in the code.

#### Basic reproduction number

The expected number of secondary infections produced by a single infection introduced into a naive population (basic reproduction number) can be caluclated as:

$$R_{0_{x}}=\beta_{x,0}\left( \frac{p_{a}\zeta}{r_{1}}+\frac{(1-p_{a})}{\gamma_{2}}+\frac{(1-p_{a})p_{m_{x}}}{r_{2}}+\frac{(1-p_{a})(1-p_{m_{x}})}{\tau_{s}}+\frac{(1-p_{t_{x}})(1-p_{a})(1-p_{m_{x}})}{r_{8}} \right)$$

In this context, a ‘naive’ population is the population at the start of the epidemic when (a) there are no previously-infected individuals ($S_{x}\approx N_{x}$) and (b) there are no measures or practices in place that reduce the contact rate below baseline ($\delta_{x,0}=1$). We further assume that at this stage in the epidemic there will be hospital beds available for all severe and critical cases that access care ($\alpha_{1}=1$).

#### Time-varying reproduction numbers

The reproduction number is assumed to vary over time, reflecting changes in the contact rate that result from both government-enforced and individually-enacted measures. We refer to two types of time-varying reproduction numbers: $R_{c}(t)=\delta_{t}R_{0}$ denotes the hypothetical reproduction number at a given point in time that would be observed in the absence of previously-infected individuals, where $\delta_{t}$ is a proportional reduction from baseline; $R_{e}(t)=R_{c}(t)S(t)/N(t)$ denotes the realized reproduction number at a given point in time, taking into account accumulation of infection and immunity in the population.

### Findings: Projected provincial cases and resource needs in the next six months (May 2020 – Jan 2021)

Across provinces, projections of deaths and cases requiring hospitalisation are lower than our previous sets of estimates (Supplementary Figures 1-9 and Tables 4-12). Across provinces, estimates of all COVID-19 related deaths are almost double those of the reported COVID -19 related deaths occurring in hospital which the DATCOV dataset aims to capture, and the number of hospital beds that we estimated would have been needed are more than those estimated to have been used over the last weeks, with the largest difference in the Eastern Cape where more than twice as many ICU beds would have been required.

Figure 1: Projections of cases, deaths and resources needed: Eastern Cape. The red crosses in the bottom right-hand panel represents 80% of the excess deaths found in the SAMRC analysis

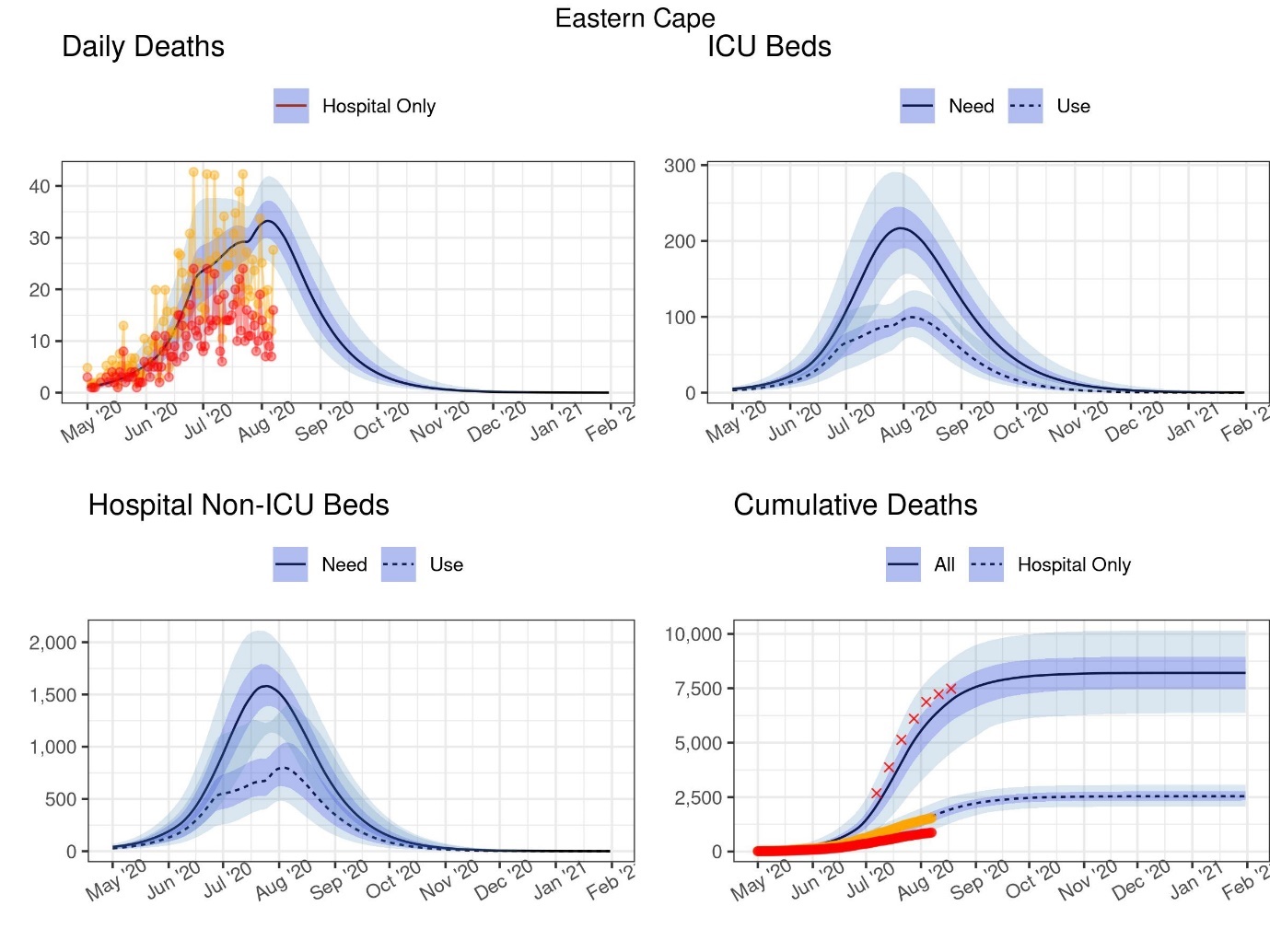

DATCOV data

Adjusted DATCOV data

Table 4: Projections of cases, deaths and resources needed at select dates: Eastern Cape

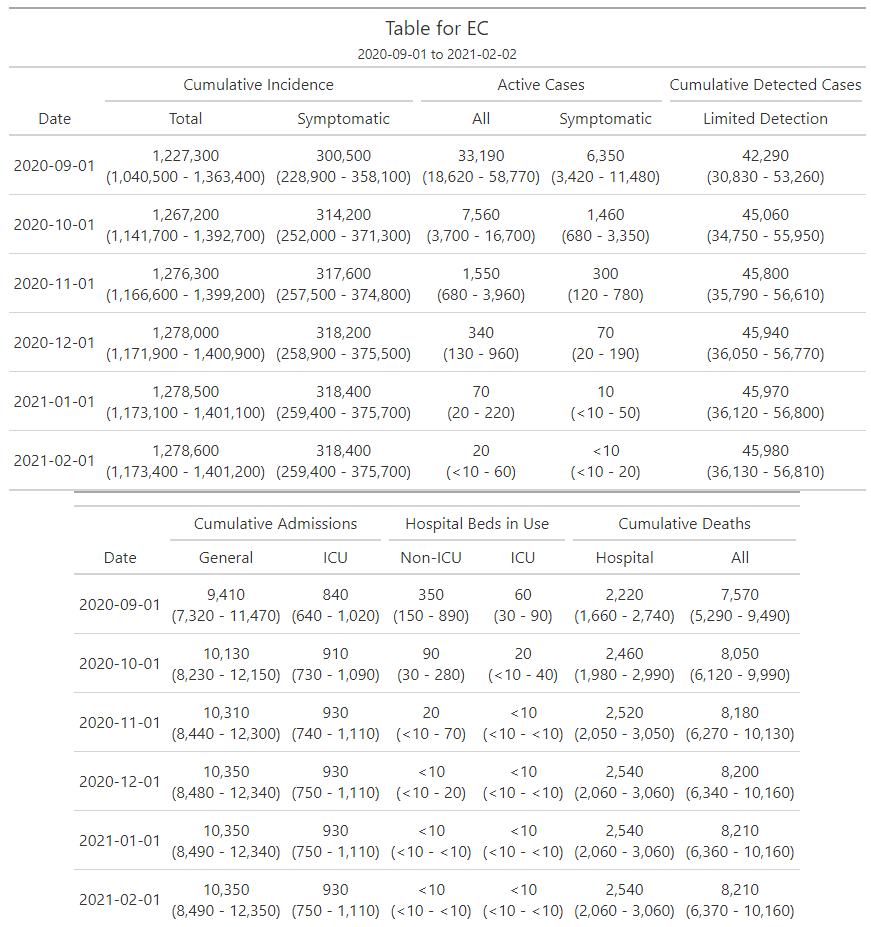

Figure 2: Projections of cases, deaths and resources needed: Free State. The red crosses in the bottom right-hand panel represents 80% of the excess deaths found in the SAMRC analysis

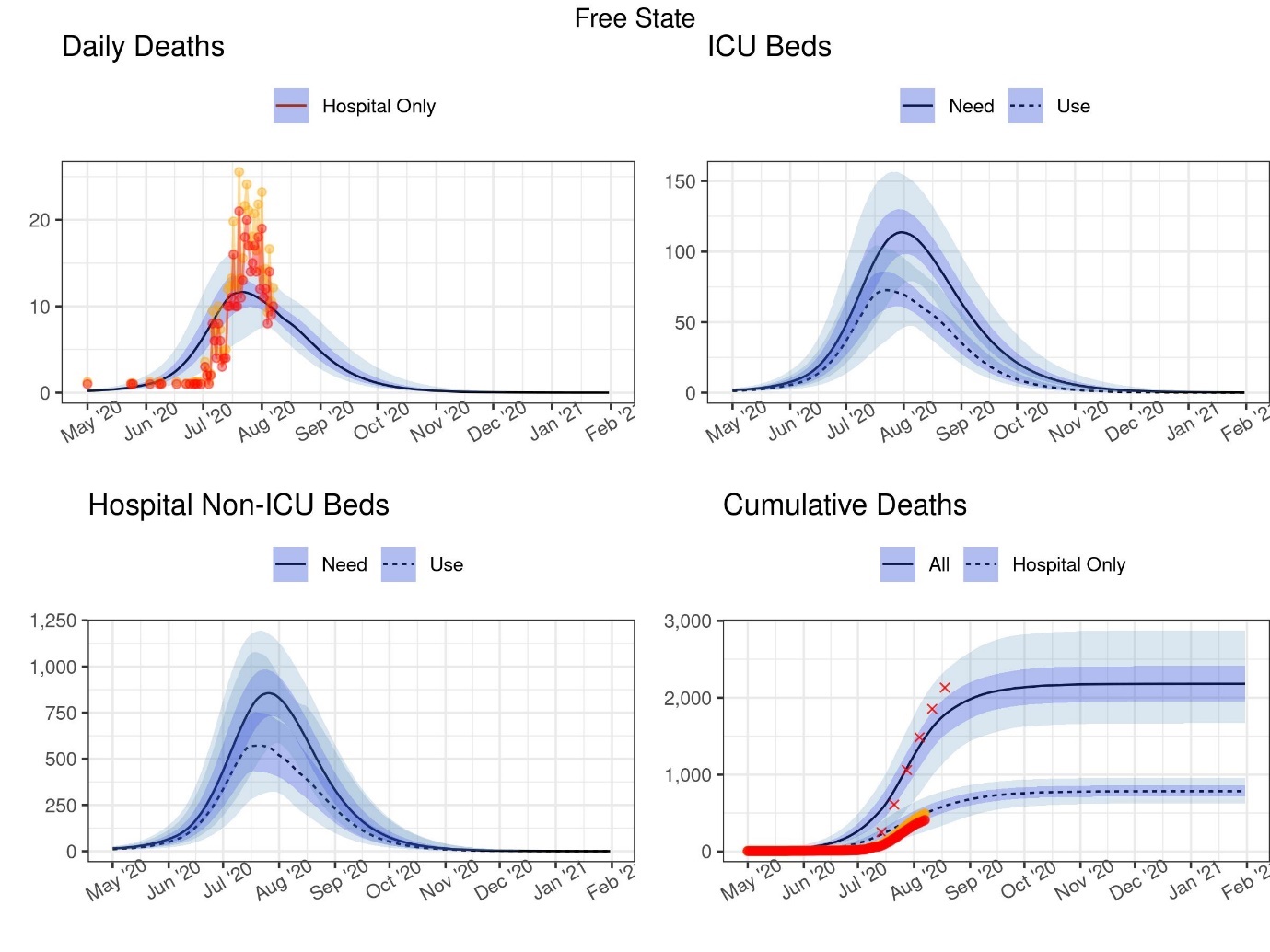

DATCOV data

Adjusted DATCOV data

Table 5: Projections of cases, deaths and resources needed at select dates: Free State

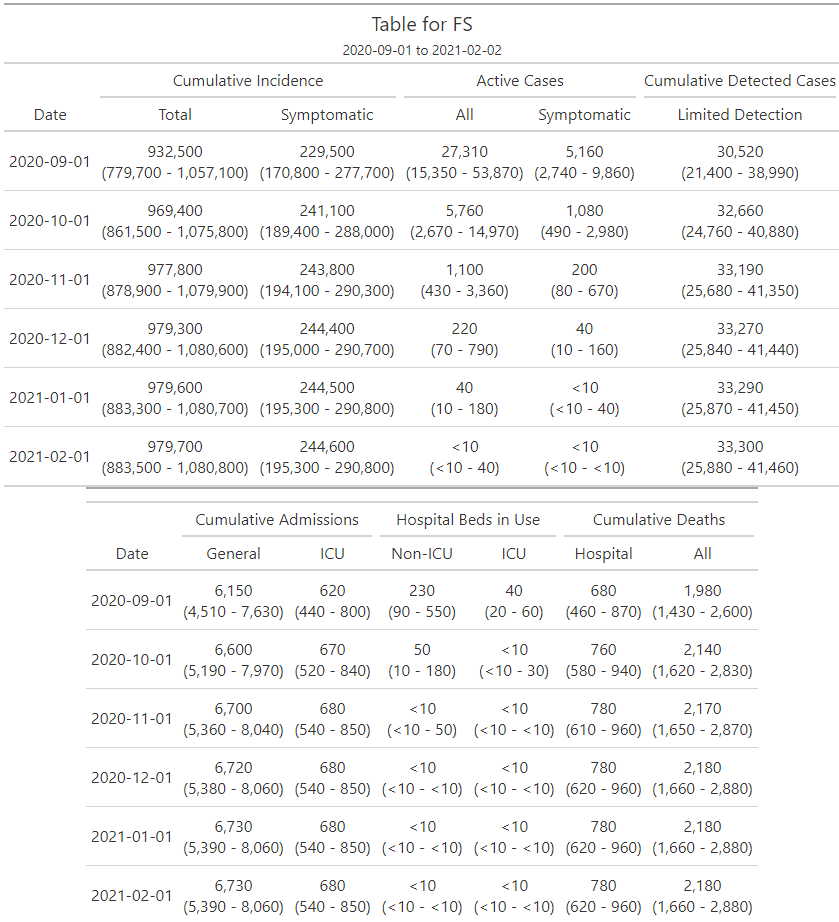

Figure 3: Projections of cases, deaths and resources needed: Gauteng. The red crosses in the bottom right-hand panel represents 80% of the excess deaths found in the SAMRC analysis

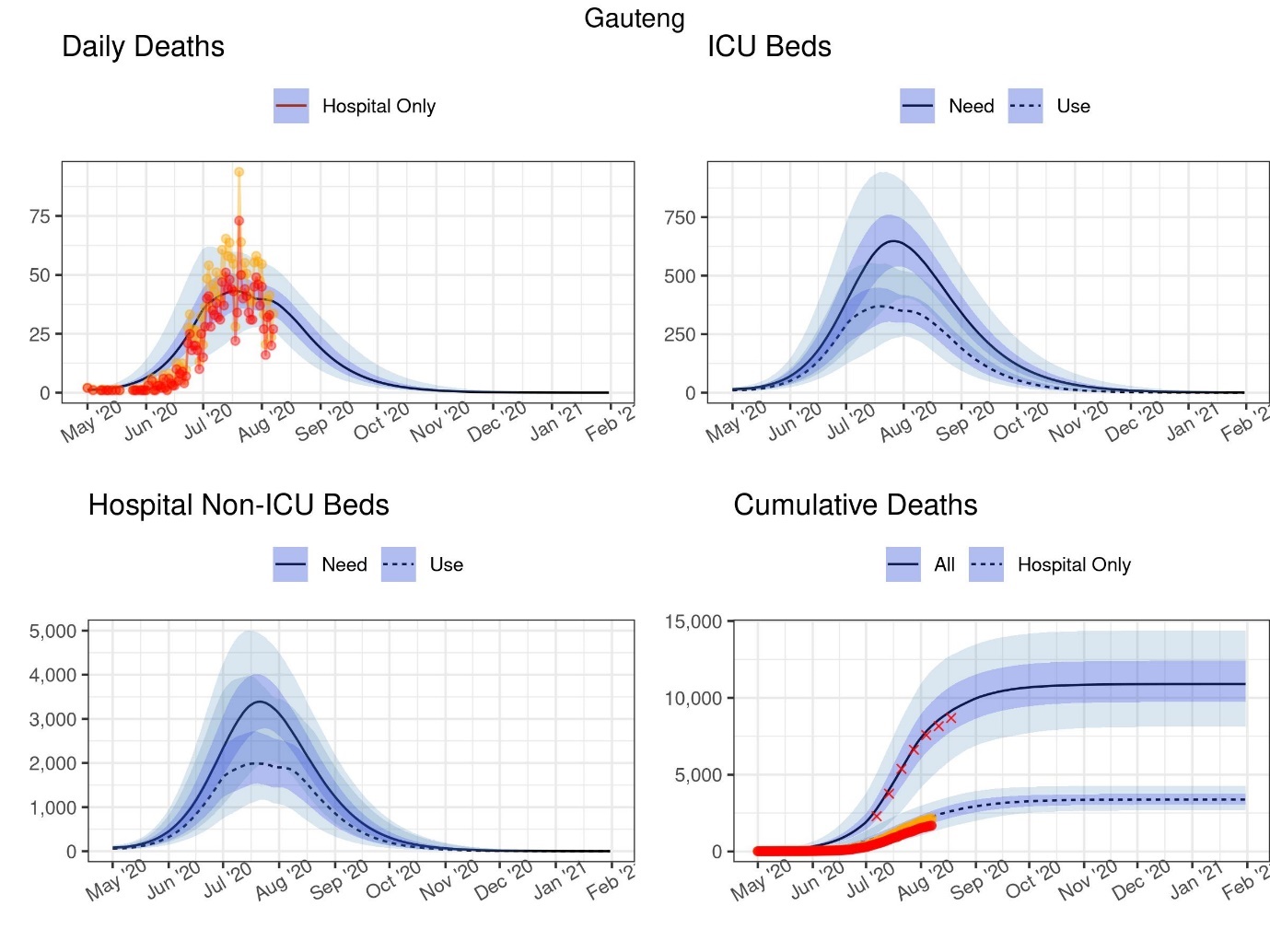

DATCOV data

Adjusted DATCOV data

Table 6: Projections of cases, deaths and resources needed at select dates: Gauteng

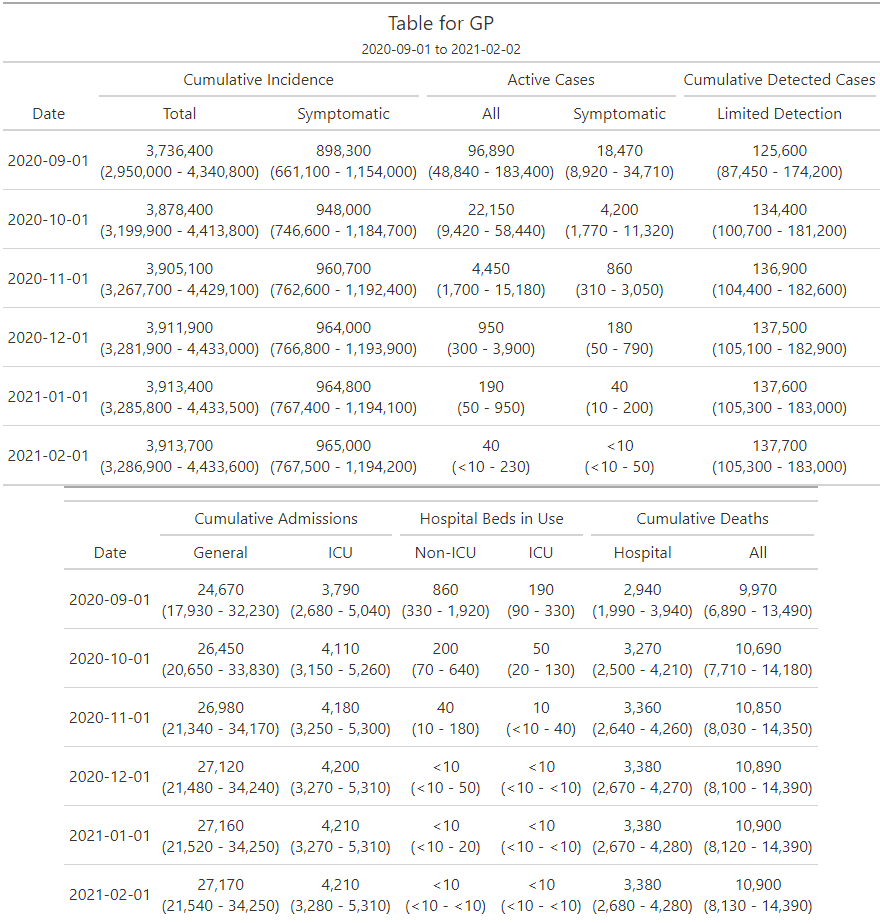

Figure 4: Projections of cases, deaths and resources needed: KwaZulu-Natal. The red crosses in the bottom right-hand panel represents 80% of the excess deaths found in the SAMRC analysis

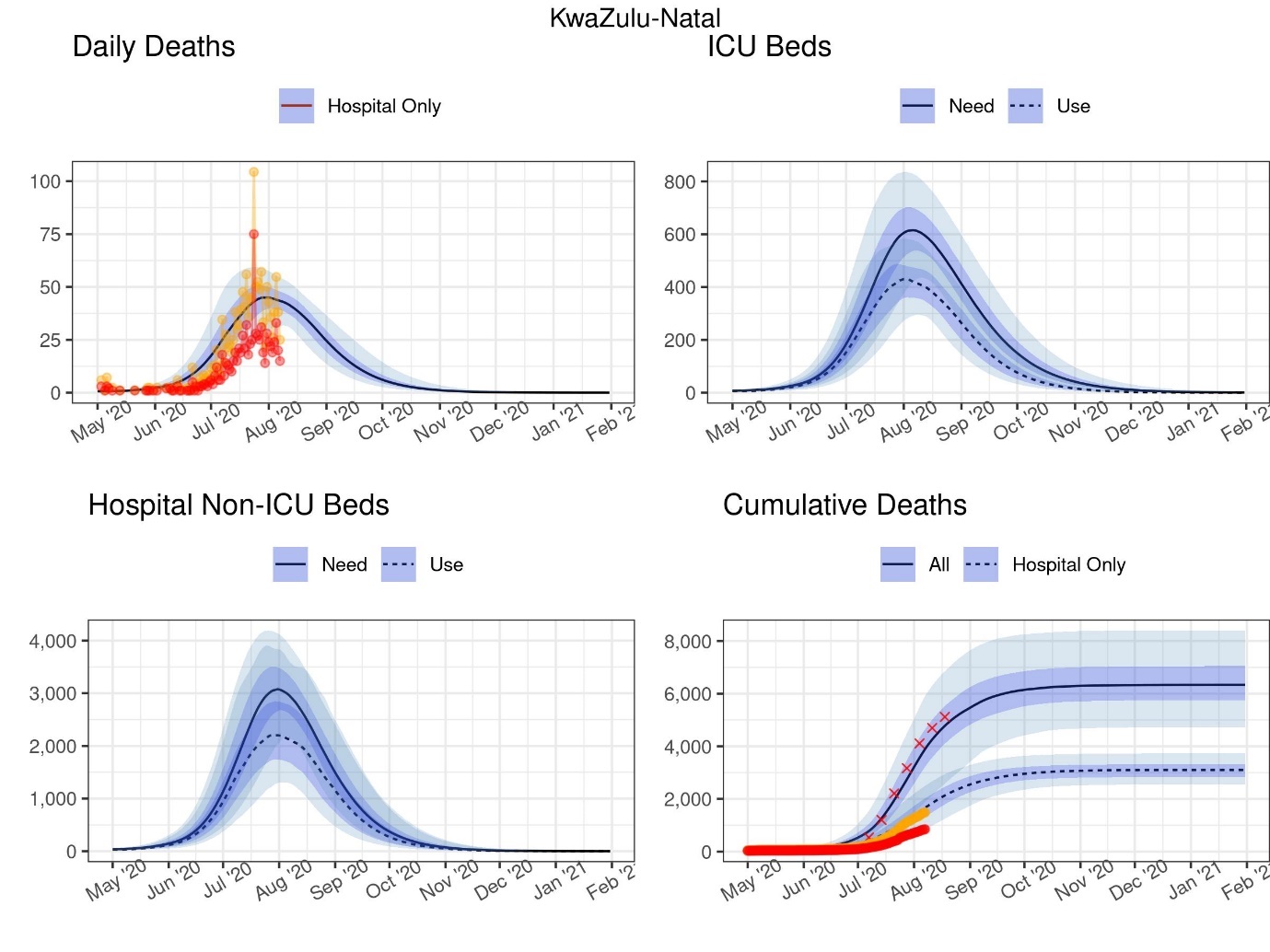

DATCOV data

Adjusted DATCOV data

Table 7: Projections of cases, deaths and resources needed at select dates: KwaZulu-Natal

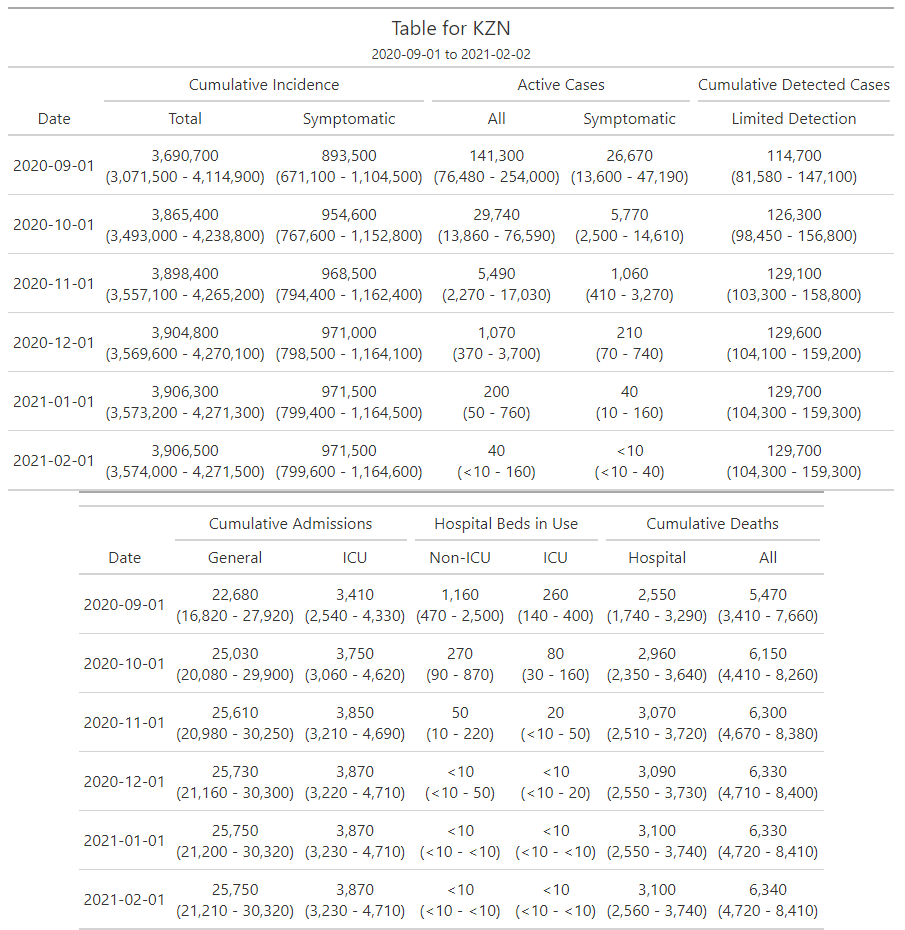

Figure 5: Projections of cases, deaths and resources needed: Limpopo. The red crosses in the bottom right-hand panel represents 80% of the excess deaths found in the SAMRC analysis

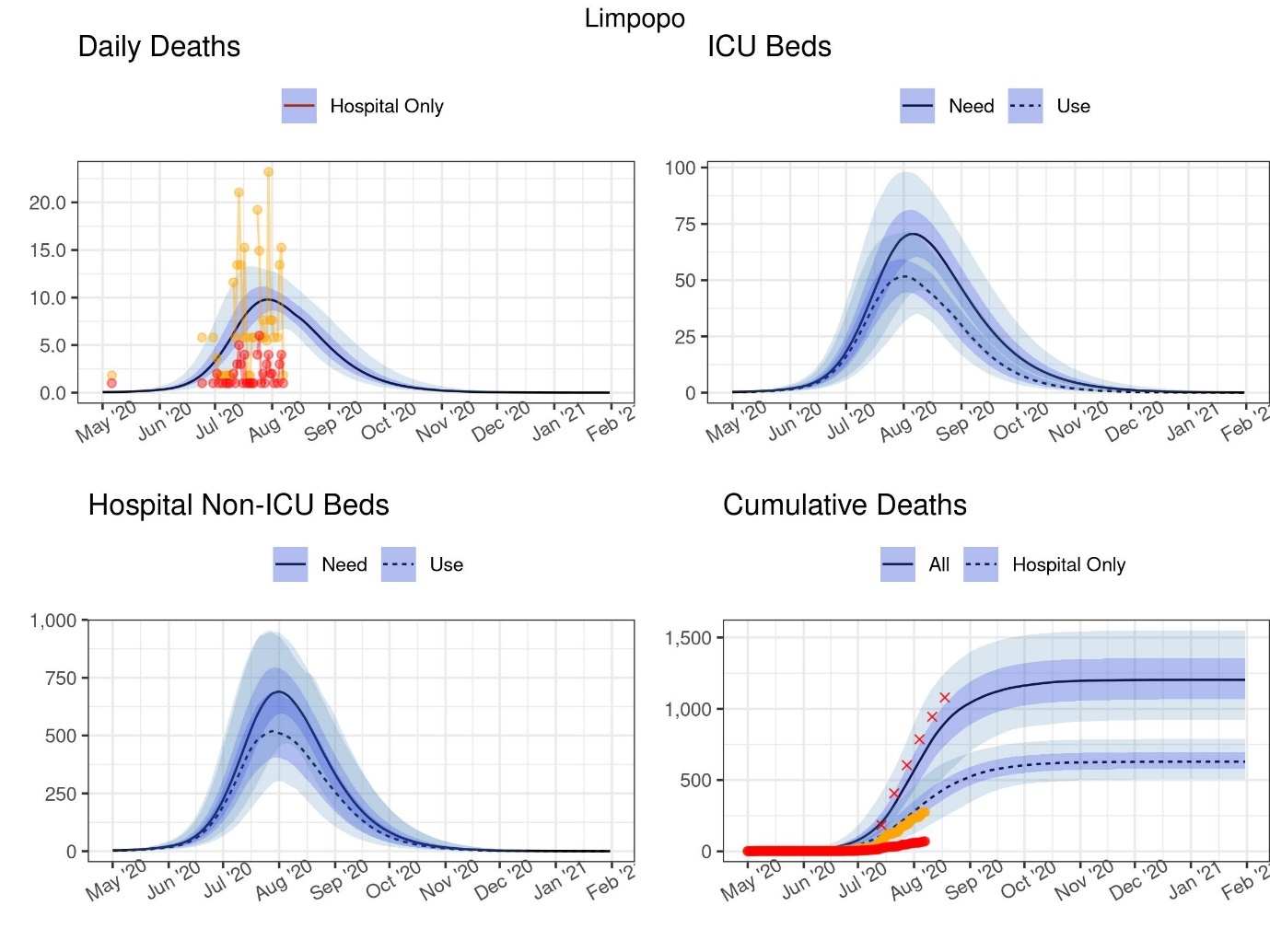

DATCOV data

Adjusted DATCOV data

Table 8: Projections of cases, deaths and resources needed at select dates: Limpopo

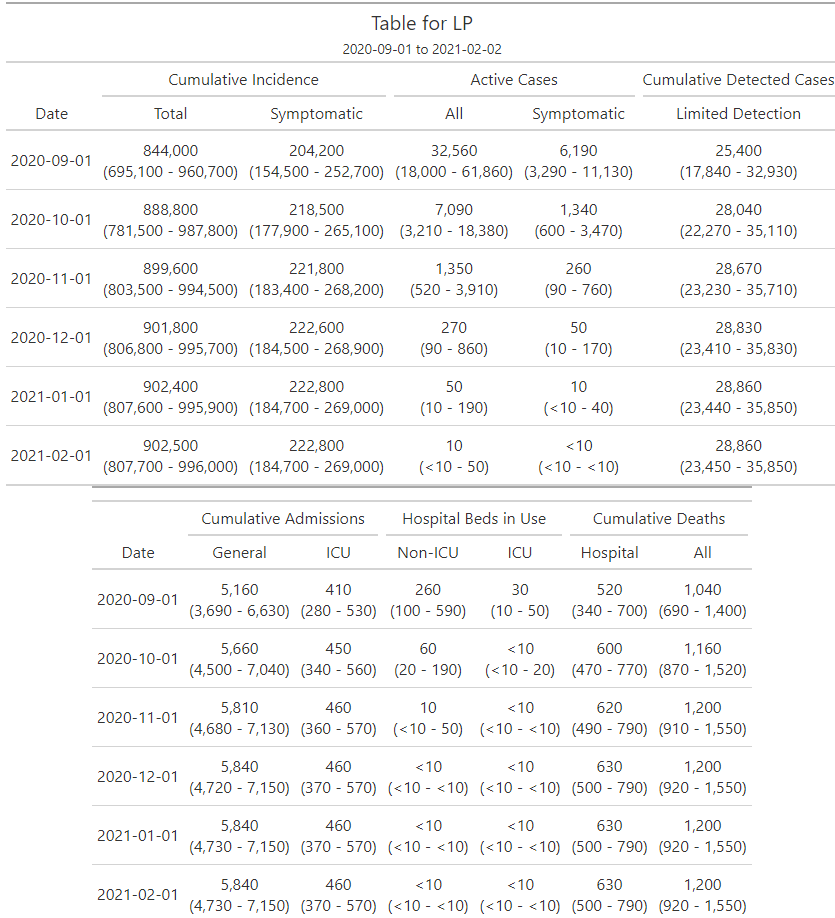

Figure 6: Projections of cases, deaths and resources needed: Mpumalanga. The red crosses in the bottom right-hand panel represents 80% of the excess deaths found in the SAMRC analysis

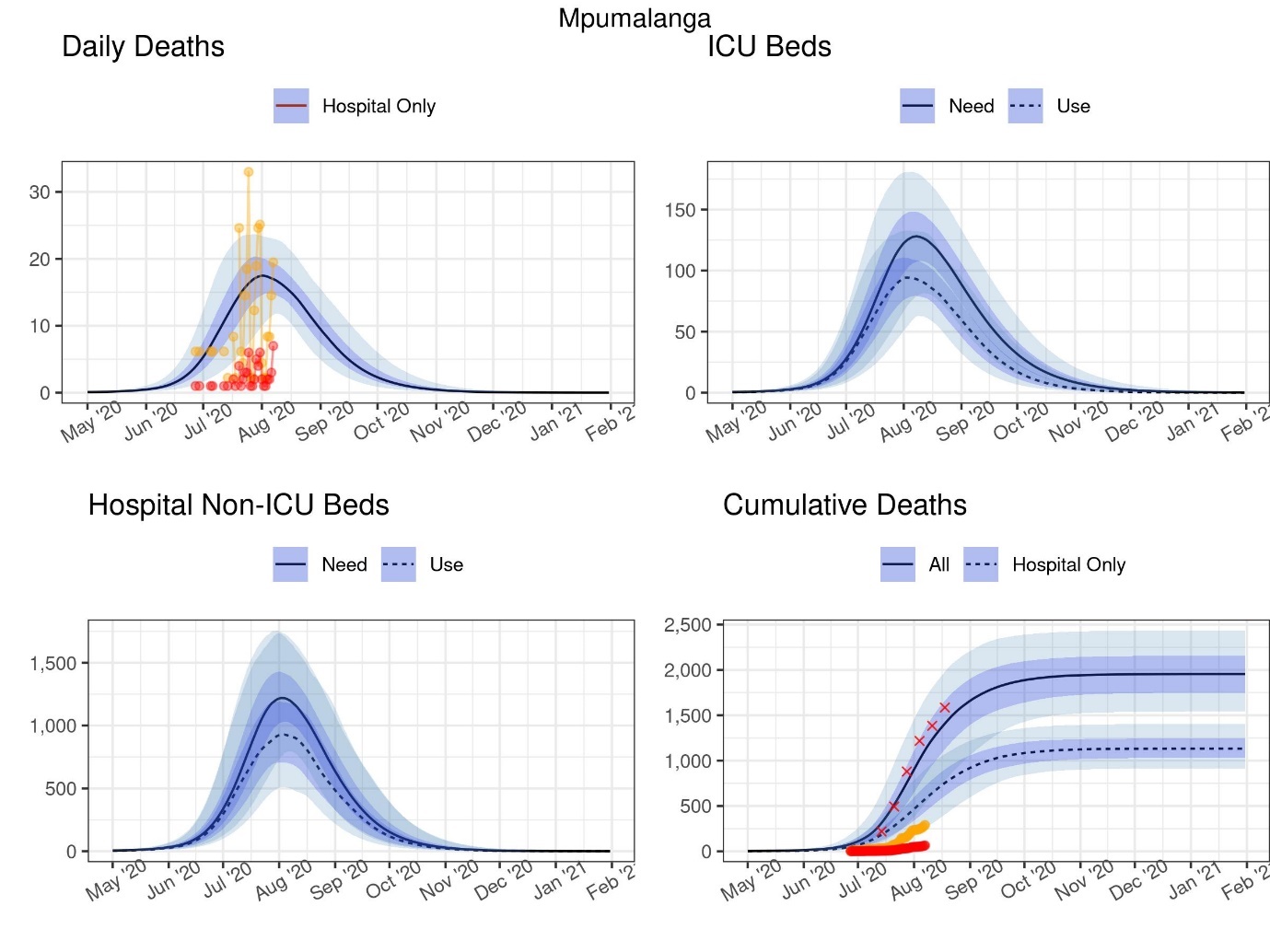

DATCOV data

Adjusted DATCOV data

Table 9: Projections of cases, deaths and resources needed at select dates: Mpumalanga

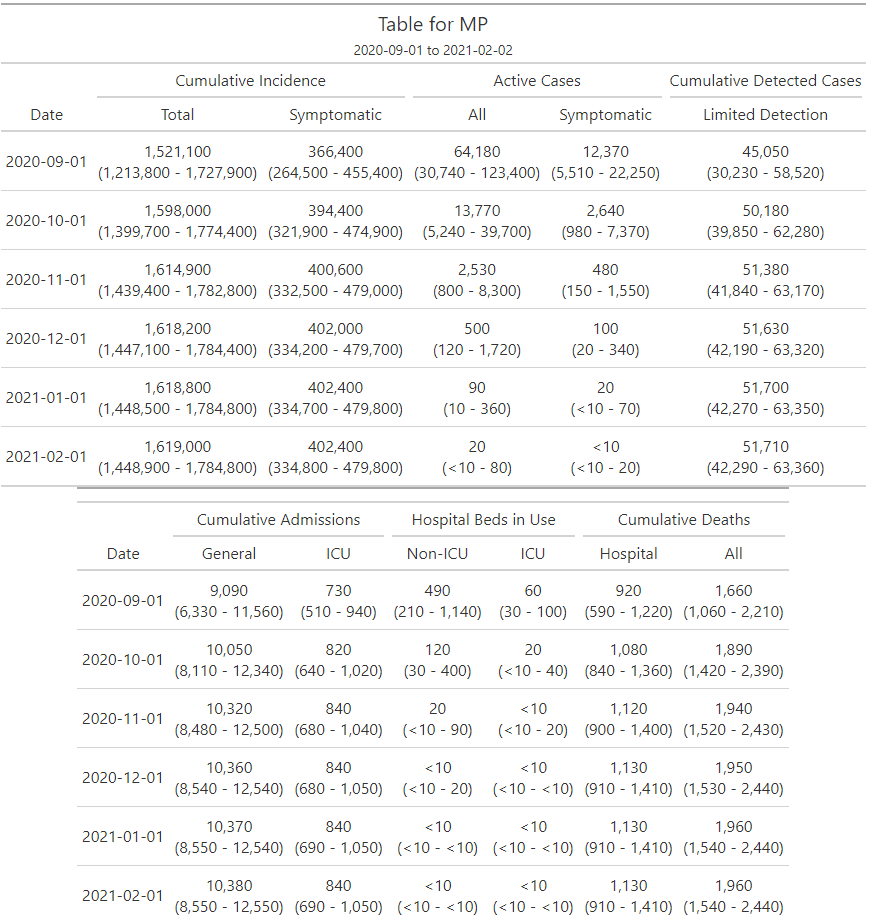

Figure 7: Projections of cases, deaths and resources needed: Northern Cape. The red crosses in the bottom right-hand panel represents 80% of the excess deaths found in the SAMRC analysis

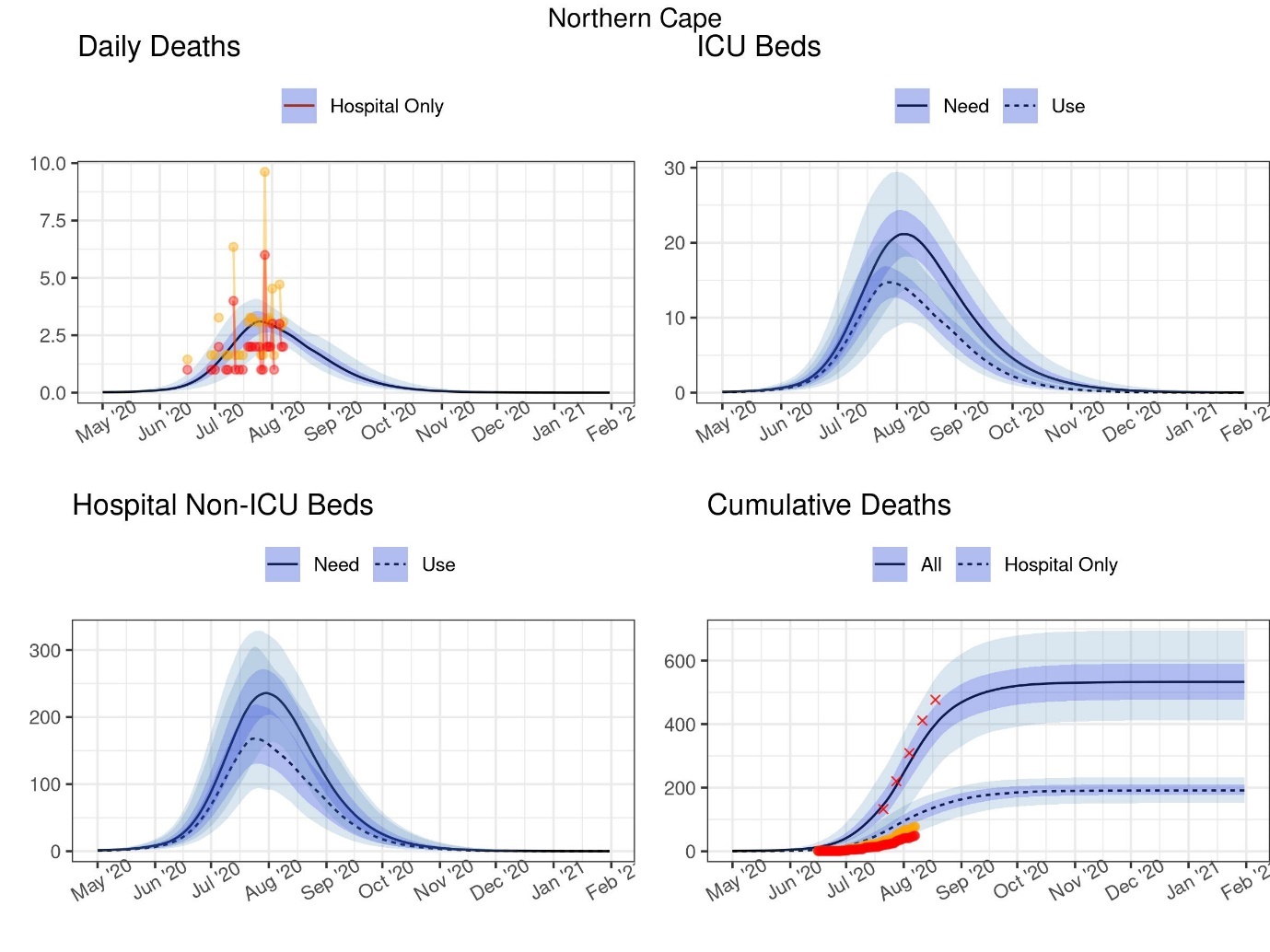

DATCOV data

Adjusted DATCOV data

Table 10: Projections of cases, deaths and resources needed at select dates: Northern Cape

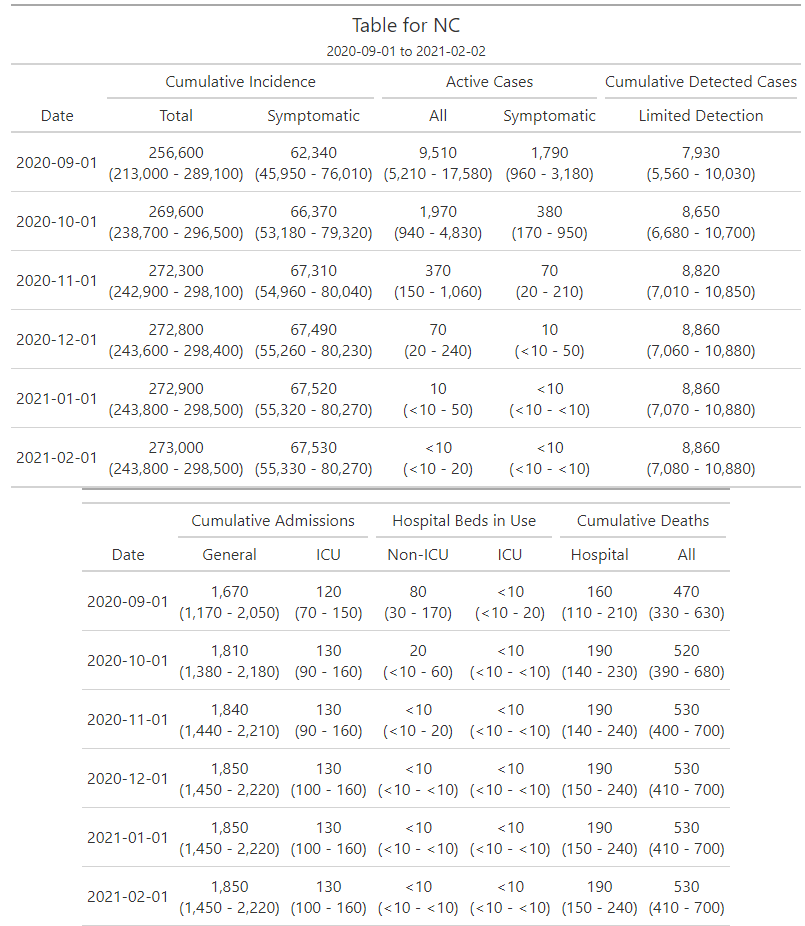

Figure 8: Projections of cases, deaths and resources needed: North West. The red crosses in the bottom right-hand panel represents 80% of the excess deaths found in the SAMRC analysis

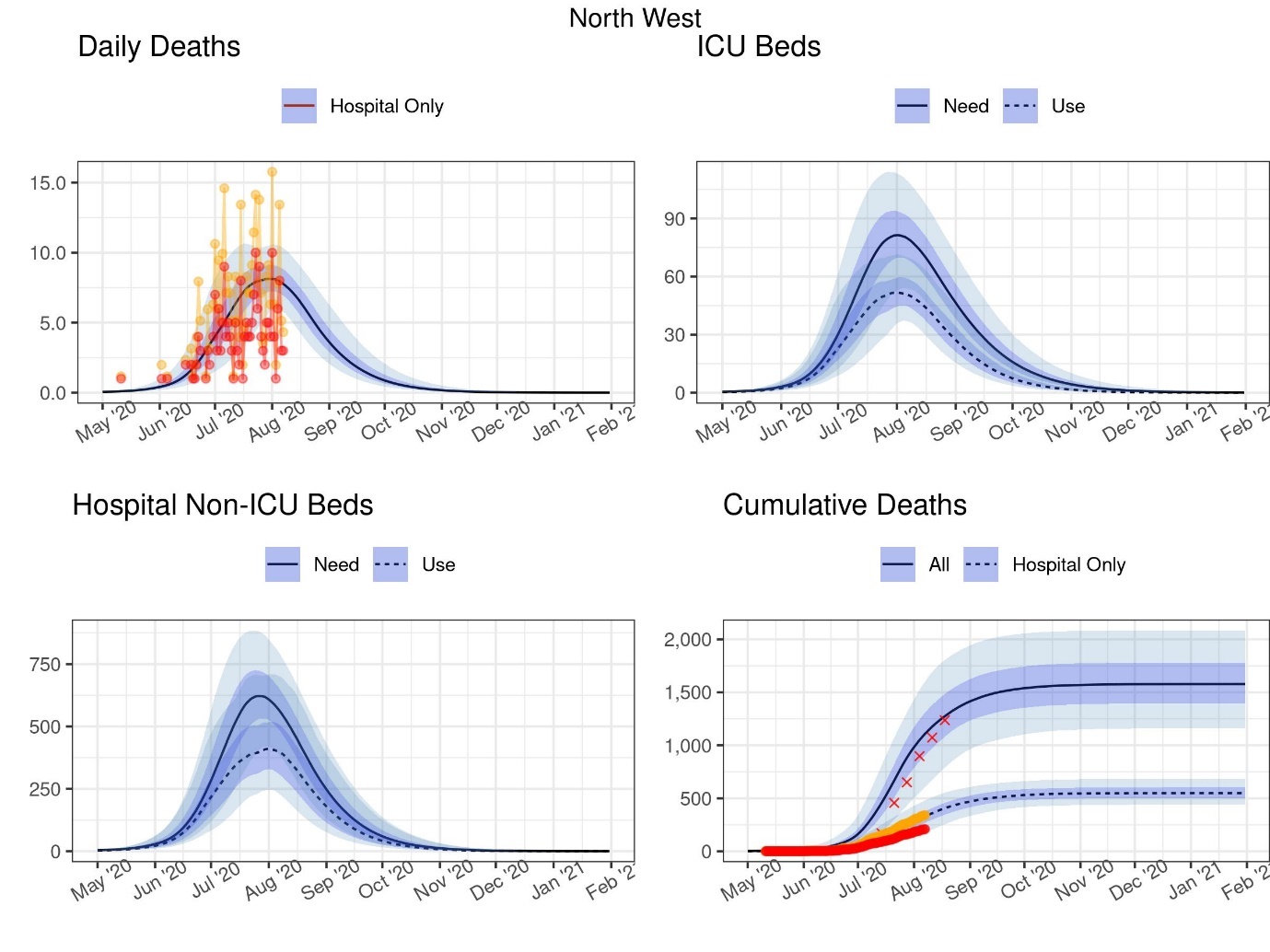

DATCOV data

Adjusted DATCOV data

Table 11: Projections of cases, deaths and resources needed at select dates: North West

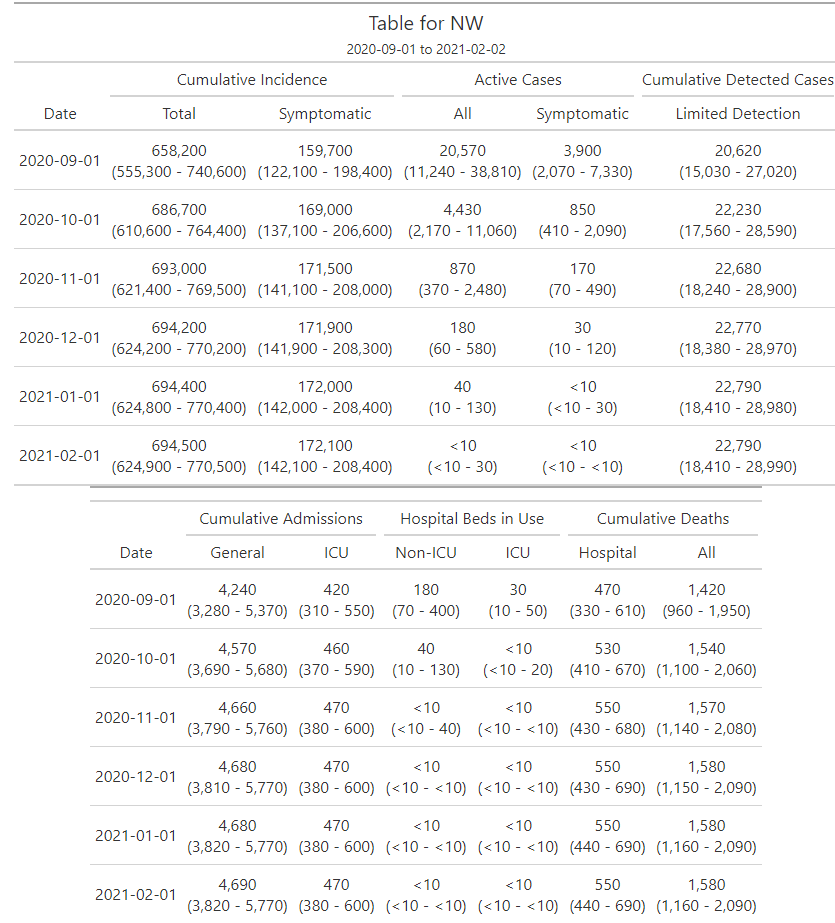

Figure 9: Projections of cases, deaths and resources needed: Western Cape. The red crosses in the bottom right-hand panel represents 80% of the excess deaths found in the SAMRC analysis

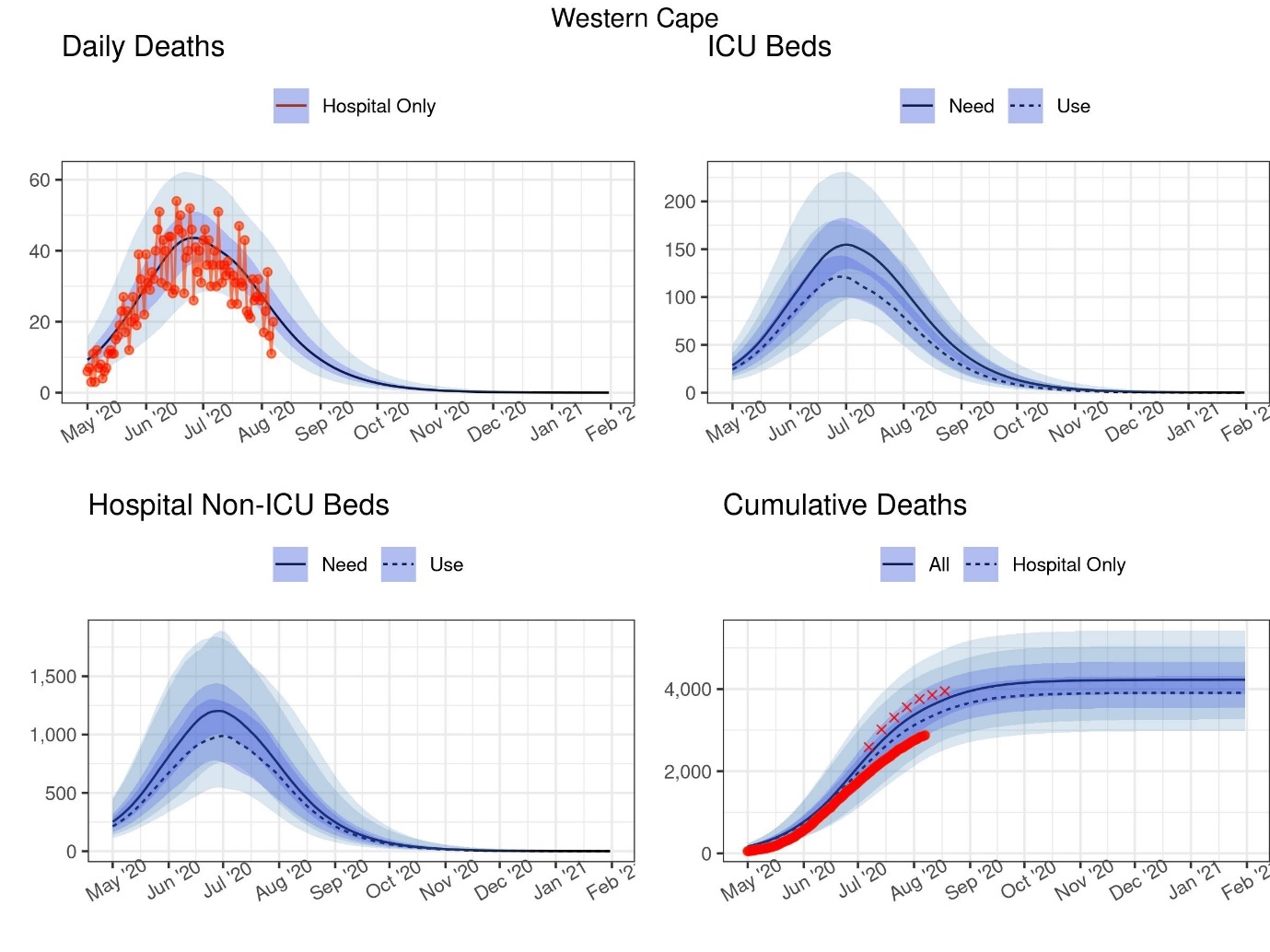

DATCOV data

Adjusted DATCOV data

Table 12: Projections of cases, deaths and resources needed at select dates: Western Cape

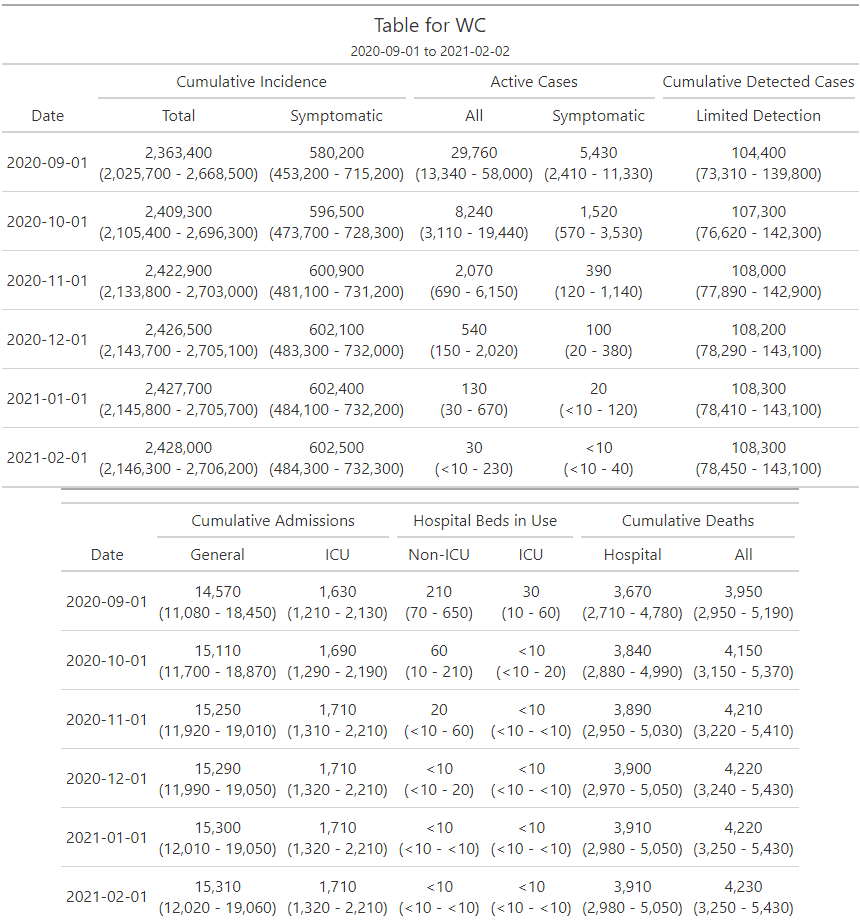

This work is licensed under a Creative Commons Attribution 4.0 International License.
